# Analysing Spatial and Temporal Concept Drift in Forecasting Malnutrition Admissions in Kenya

**DOI:** 10.64898/2026.11.18.26367901

**Authors:** Steven Squires, Tinkle Chugh, Theodoros Economou, Peter Hailey, Matt Fortnam

**Affiliations:** Department of Computer Science, University of Exeter, Exeter, UK; Department of Mathematics and Statistics, University of Exeter, Exeter, UK; Centre for Humanitarian Change, Nairobi, Kenya; Agile Earth, Cornwall, UK; Environment and Sustainability Institute, University of Exeter, UK

**Keywords:** machine learning, spatial heterogeneity, spatial non-stationarity, data drift, temporal non-stationarity, spatial drift, malnutrition data, healthcare surges, Kenya

## Abstract

Forecasting malnutrition admissions could support anticipatory planning in health systems, but existing machine learning and statistical approaches have struggled to produce forecasts of practical value. We investigate whether this disappointing performance is partly attributable to spatial and temporal concept drift, whereby relationships between predictors and admissions differ by location and over time, respectively. We also examine whether adaptations to the modelling approach can mitigate these effects. We used malnutrition admissions from 44 subcounties across 14 counties in Kenya between 2015 and 2020, alongside potential predictors including normalized difference vegetation index (NDVI) and rainfall. We conducted univariate, multivariate, and non-linear analyses of spatial and temporal concept drift, and then trained machine learning (ML) forecasting models using adaptations intended to mitigate these drifts. We find substantial spatial and temporal variation in relationships between predictors and admissions. For temporal concept drift in one subcounty the maximum correlation between admissions and time-lagged admissions for different time periods ranged from −0.39 (95% CIs: −0.74 to −0.04) to 0.45 (95% CIs: 0.11 to 0.79); the equivalents for changes between subcounties ranged from a minimum to maximum of −0.18 (95% CIs: −0.45 to 0.09) to 0.64 (95% CIs: 0.43 to 0.85). For non-linear models, there are also substantial changes in feature importance. At a forecast horizon of three months, our best-performing ML model produced a root mean squared error of 1.07 (95% CIs: 0.96 to 1.18), significantly outperforming the best baseline of 1.23 (95% CIs: 1.10 to 1.39); however, the predictions are unlikely to be sufficiently accurate for practical use. These findings suggest that spatial and temporal concept drift should be explicitly considered when developing forecasting models for malnutrition admissions, rather than relying directly on off-the-shelf ML approaches.

## 1 Background

Malnutrition is a serious condition [1], with an estimated 43 million children suffering from wasting [2], that affects individuals and communities and places additional pressure on health systems. Accurate forecasts of malnutrition admissions could support anticipatory action, including prepositioning supplies, adjusting healthcare staffing and seeking additional assistance [3, 4]. However, forecasting admissions remains difficult. Existing machine-learning (ML) work using Kenyan admissions data has reported limited practical performance [5], and the wider literature provides little evidence that current approaches can generate consistently actionable forecasts [6].

Part of the challenge may lie in the complex relationship between the number of people experiencing malnutrition and those subsequently admitted to health facilities linked to challenges accessing and using services as well as the ability of a health system to provide those services [7–9]. However, forecasting population malnutrition itself has also proved difficult. Agent-based approaches have struggled to outperform baselines [10], decision-tree methods have achieved only modest gains over bench-marks [11], and models using high-frequency data have shown limited correlation with observed outcomes [12]. This suggests that the difficulty is not solely the mapping from malnutrition prevalence to admissions.

A consistent finding across studies is that time-lagged measures of malnutrition or admissions tend to dominate model predictions [5, 13]. Adding exogenous variables often provides little improvement: models using lagged malnutrition prevalence alone have performed similarly to models including additional predictors [12], with comparable findings for admissions forecasting in Kenya [5]. This is notable because climatic, environmental and socioeconomic variables are known to be associated with malnutrition [14]. Understanding why these relationships do not translate into stronger forecasts may therefore be important for improving predictive performance.

A standard route to improving the value gained by using ML models is to increase the quantity of training data. For malnutrition forecasting, this generally requires adding observations from earlier periods, other geographical areas, or both. Our hypothesis is that the value of this additional data is constrained by *concept drift*. Spatial concept drift occurs when relationships between predictors and admissions differ between locations; temporal concept drift occurs when these relationships change within a location over time. If either is substantial, observations added from other places or earlier periods may not represent the relationships required for the target location and forecast date. Consequently, simply increasing the amount of training data may provide smaller gains than expected and, in some circumstances, may reduce generalisation performance.

The main contribution of this paper is to quantify spatial and temporal concept drift in malnutrition admissions data from Kenya and assess whether it provides a plausible explanation for the limited performance of current forecasting approaches. We examine drift using univariate and multivariable regression and a non-linear ML model, and then test pragmatic adaptations intended to mitigate it, including ongoing model retraining and alternative spatial pooling strategies. Our aim is not to establish concept drift as the sole cause of poor forecasting performance, but to determine whether it is sufficiently important that future forecasting approaches should account for it explicitly.

## 2 Methodology

### 2.1 Spatial and temporal concept drift

Commonly used terms for (or similar to) temporal concept drift are non-stationarity and data drift; for spatial concept drift, equivalents such as spatial non-stationarity, spatial drift, or spatial heterogeneity are often used. However, there is substantial variation in terminology; therefore, we begin by defining how we use these terms in this paper. Critically, we are referring to changes in the *patterns* between inputs and outputs with space or time, not to variation in the data itself.

We define **X**_*i*_ *∈* ℝ^*n×p*^ and **y**_*i*_ *∈* ℝ^*n*^ for region, *i* with *n* time periods (e.g., monthly or yearly); an element of **y**_*i*_ is the number of malnutrition admissions in that region for that time period and a row of **X**_*i*_ is a vector of *p* input features corresponding to that time period. We assume there exists some mapping such that *f* (**X**_*i*_) *≈* **y**_*i*_; if spatial concept drift is a problem then for a different region, *j, f* (**X**_*j*_) *≉* **y**_*j*_ and instead a different function *g* is required such that *g*(**X**_*j*_) *≈* **y**_*j*_. Similarly, for temporal concept drift we can define a desired mapping at time *t* = *t*_0_ as *f*_*t*0_ which works appropriately at that time point but that at *t* = *t*_0_ + Δ*t* does not hold, therefore *f*_*t*0_ ≠ *f*_*t*0+Δ*t*_. We will refer to “concept drift” for brevity when referring to either or both of spatial and temporal concept drift.

Learning potentially complex and noisy relationships requires sufficient data, yet expanding the dataset creates a trade-off. Adding older observations increases exposure to temporal drift, while pooling locations increases exposure to spatial drift. We therefore assess both forms of drift before testing approaches intended to mitigate them.

### 2.2 Data and preprocessing

In this paper, we use malnutrition admission data from Kenya from 2015-2020; data is available per subcounty and per month therefore each datapoint is the number of malnutrition admissions per subcounty-month. There are data on moderate acute malnutrition and severe acute malnutrition which we sum together, as is common practice [5, 12], we also sum across numbers of outpatients and inpatients. There are around 300 subcounties within Kenya but we exclude all those where there is missing data leaving us with data from a total of 44 subcounties from 14 counties. In the left plot of Figure 1 we show a map of Kenya with the 44 included subcounties, the colours and labels represent the 14 counties; in the right plot we show labelled subcounties with available data from two counties (Marsabit and Turkana) which are part of Kenya’s arid and semi-arid lands (ASAL) which often have significant issues with malnutrition [15].

**Fig. 1.**
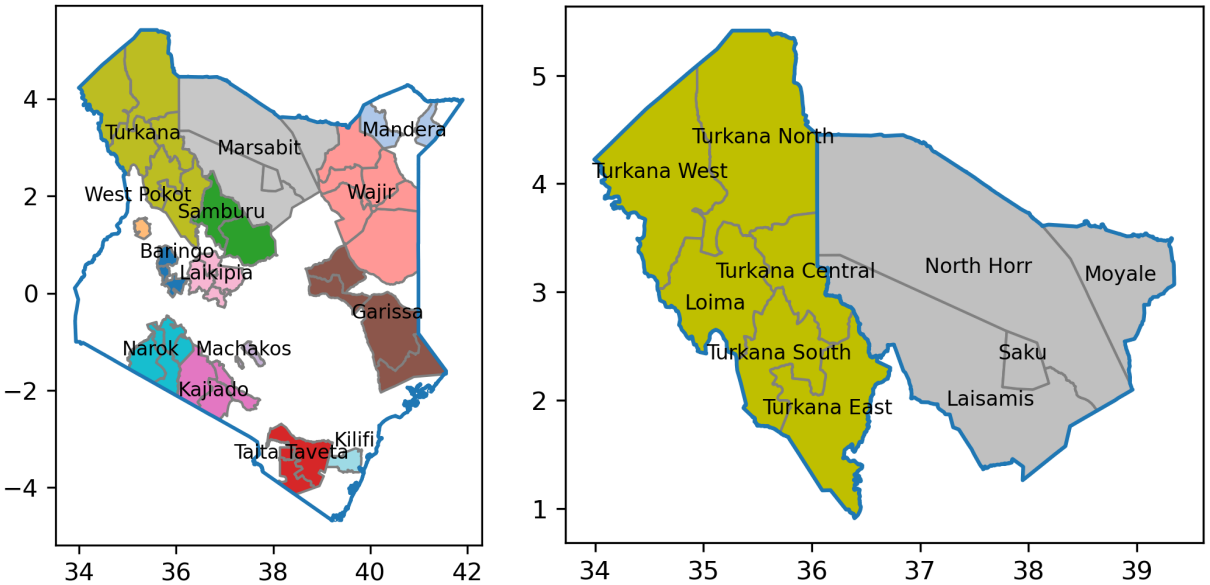
Left) Map of locations of admissions data used within Kenya. The labels show the counties; the colors are by county but shown only for those subcounties where we have data. Right) Map of two counties (Marsabit and Turkana) with subcounties labelled.

As input data, we use variables primarily from publicly available sources and generally believed to be being associated with malnutrition: time-lagged values of the malnutrition admissions, diarrhoea admissions, Integrated Food Security Phase Classification (IPC) estimates from FEWS NET, diesel prices, total rainfall, minimum and maximum temperature, normalised difference vegetation index (NDVI), malaria admissions, maize meal prices, number of conflict events and fatalities. Details of these input variables, where they were retrieved from, and details on preprocessing are in the supplementary material as text and in supplementary Table S1.

All input data are standardised to z-scores within each subcounty by subtracting the mean and dividing by the standard deviation. For rainfall, temperatures and NDVI, we standardise by month as well as subcounty. For assessment of statistical relationships between variables and, in particular, for assessing spatial and temporal concept drift, we utilise the whole dataset, standardising across all time. When training models to perform forecasting, we split into a training and testing set by time (testing set data from later dates) with a cutoff at 2019.

With regard to training the models, we utilise two approaches, one standard and one via ongoing training (see Section 2.4); for the standard approach the data is standardised on the training set alone with the testing set then standardised using the mean and standard deviations from the training set so there is no danger of data leakage; for ongoing training we re-standardise the data for each model trained.

We used forecasting horizons (number of months forward we are making predictions at) for admissions at 1 and 3 months. When training forecasting models we utilised input variables with time lags of 0, 1, 2 months.

### 2.3 Exploring concept drift

If concept drift is a significant issue it should manifest in apparent patterns in feature importance as a function of space or time. To explore this we consider three approaches: a univariate regression (also called simple linear regression), a multi-variable regression, and a gradient boosted decision tree, XGBoost [16]. The purpose of including multi-variable regression is that adding covariates (additional inputs) to the model will alter the coefficient of the variable of interest and should therefore be assessed. The reason for exploring the XGBoost regressor is that this is a widely used method for predicting malnutrition and provides non-linear and interaction terms into the predictions.

For the univariate and multi-variable regression we consider the coefficient between the malnutrition admissions and the input variable; for univariate regression as the data is standardised, these coefficients are identical with the Pearson correlation coefficient between the input and output. XGBoost implements gradient boosted decision trees so we cannot directly assess coefficients, instead we perform feature selection using cross-validation, reporting which features are included at the end of the feature selection process.

For spatial concept drift we consider the effect on features when training on individual subcounties, counties, livelihood zones (see supplementary material and Figure S1 for details), and across all the subcounties combined. For spatial concept drift the key question is whether the variables have similar importance (coefficients for linear regression or retained features for XGBoost) across regions in a structured way (say similarity in neighbouring regions). If the feature importance is similar for all regions then a plausible model is one where the features affect the admissions in the same way independent of region. For temporal concept drift the question is whether the importance of features is consistent across time for the same subcounty, or grouping of subcounties; we use sliding time windows of 30 months to train the models and study how coefficient values, or retained features, change with time.

### 2.4 Approaches to mitigate concept drift

We will provide evidence that suggests that spatial and temporal concept drift may be significant problems, therefore we also explore approaches to mitigate concept drift. In standard machine learning the training, validation and test sets are split once, and for time-series it is common to split them temporally - with training data being furthest back in time, with validation data following and testing data being closest to the present. But if temporal concept drift is a significant problem then training on data from further back in time is likely to result in the wrong patterns being learned; to mitigate this we utilise ongoing training where a new model is trained for every time point (month) throughout the test set. Our test set is data from 2019, for standard training we train the model on all the training/validation data (from 2015 to 2018 inclusive). For ongoing training we train each model using all data prior to the test date - meaning that each model is only used to predict one time-horizon forwards. By doing so we maximise the quantity of data we have and reduce the effect of temporal concept drift by incorporating as much contemporary data as is possible; this also dilutes the effect of data from further back in time.

For spatial concept drift our approach is to train on different groups of subcounties, assessing whether we can improve performance by doing so. We therefore train models: individually on a subcounty; together on all the subcounties within a county; together for all the subcounties within a defined livelihood zone; together for all available subcounties. The models trained on the individual subcounty will have the least spatial concept drift but suffer from the smallest quantity of data. For the county, livelihood and all combined training approaches we increase the quantity of data but at the expense of potential spatial concept drift.

### 2.5 Models, training and baselines

For the forecasting models we used the elastic net [17] and an XGBoost [16] regressor. The elastic net is a regularised linear regression model which combines *L*_1_ (Lasso) [18] and *L*_2_ regression (ridge/Tikhonv) [19]. XGBoost is a commonly used method for tabular data [20, 21] and regularly used for malnutrition forecasting [5, 13, 22] which allows for both interaction terms between input variables as well as non-linear functions to be utilised. We trained models on all the input data and also on just the time-lagged values of the admissions.

We utilise several plausible baseline measures to compare the model predictions against: the value of the last output, the average of the last three outputs, the average admissions, and the average admissions for that month. For all the baselines we also perform the forecast at the four different spatial scales: all subcounties, individual subcounties, by county, and by livelihood zone.

### 2.6 Statistical analysis

The comparisons were made at the subcounty level or concatenated across all sub-counties, regardless of the approach used for making the forecast; we used root mean squared error (RMSE) between the outputs and the forecasts. For metrics we quantified uncertainties by bootstrapping the results and present them as 95% confidence intervals. For all coefficients of the linear regressions uncertainties are calculated using classical standard errors and are reported at the 95% level. For training XGBoost we used scikit-learn [23]; for the linear regressions we used a combination of scikit-learn and statsmodels [24].

## 3 Results

### 3.1 Admissions data

Admissions data varies substantially between subcounties both in absolute values, and relative changes in magnitude; we show examples of absolute numbers of admissions for the four subcounties of Marsabit county in Figure 2. We standardised the admissions by subcounty in all our analyses, with plots of the standardised admissions for all the counties shown in supplementary Figures S2 to S15. The correlation for admissions between subcounties is highly variable with correlations rarely above ~0.5, in supplementary Figure S16 we show correlations between all subcounties. The admissions data shows substantial fluctuations, the number of admissions often alters considerably over as little a period as one month, and this appears to occur both with rising and falling admissions. As an example for North Horr (Figure 2) in April 2018 admissions were 269, for May 2018 they were 1,279, and for June 2018 they were 273.

**Fig. 2.**
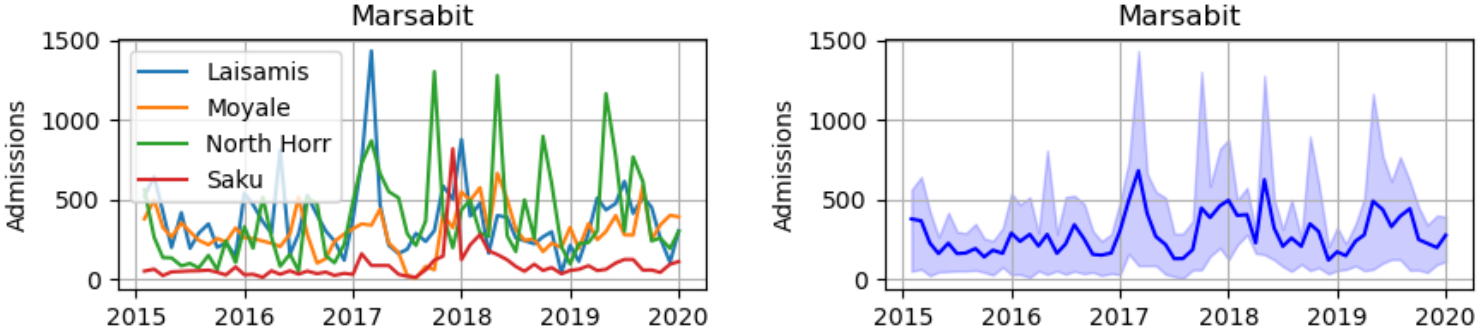
Example admissions data from the county Marsabit. Left) the number of malnutrition admissions per month for the four subcounties making up Marsabit county (Laisamis, Moyale, North Horr, and Saku). Right) The mean (blue line) value of admissions of the subcounties per month with the range of the admissions shown as blue shading.

### 3.2 Spatial concept drift

In Figure 3 we show coefficients generated from univariate regression between the specified variable (admissions, FEWS NET IPC, and NDVI) and the admissions for a forecast horizon of three months. The top plot shows the coefficients for the individual 44 subcounties; we also show the coefficient if all subcounties are concatenated before regression (large circle marker and with much smaller confidence intervals). These variables are sorted separately for each variable, i.e. the ranking order is not the same for each variable. For each subcounty the uncertainties in the coefficients are high even though the number of samples is 57, giving a reasonable sample-to-feature ratio; this means the coefficients could plausibly take one of a wide range of values.

**Fig. 3.**
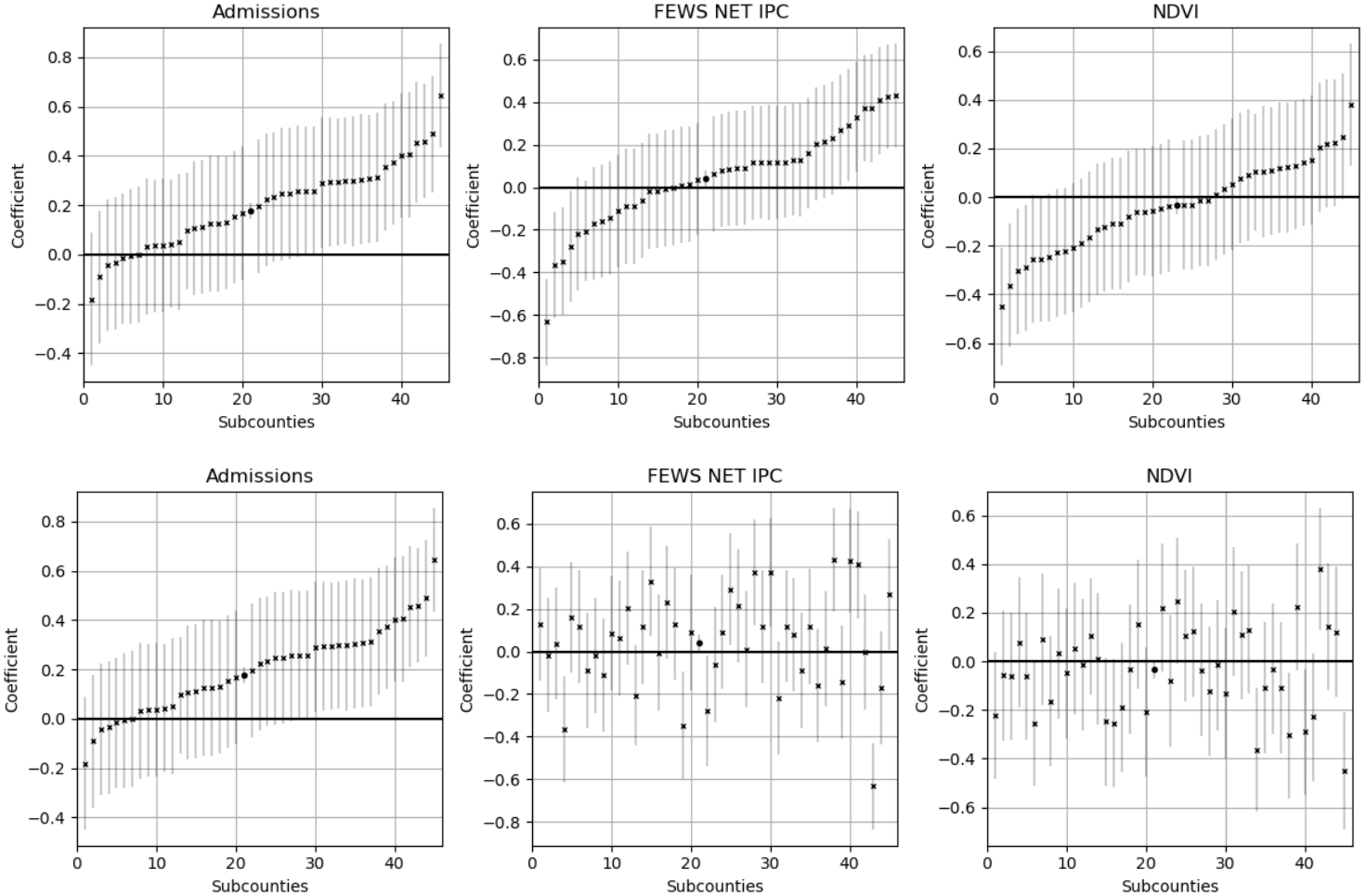
Univariate coefficients by subcounty for a forecast horizon of three months. Also shown is the coefficient if all data across the subcounties is concatenated, shown as a large circle marker and with much smaller confidence intervals. Top) The coefficients have been sorted separately for each variable to show the overall spread of the coefficients across the subcounties. Bottom) The coefficients have been sorted by the same ordering as for the admissions coefficients, to show the lack of obvious relationship if we order by the admissions coefficient.

In the bottom plot of Figure 3 we show the same coefficients (forecast horizon of three months) ordered by the ranking of the admissions coefficients, noting that in the literature the time-lagged malnutrition measure is often the most important predictor therefore models are likely to prioritise the weighting of the admissions variable. A high coefficient for the admissions does not imply in a high coefficient for the FEWSNET IPC or NDVI variables. Comparable results for the other variables are shown in supplementary Figures S17 to S19, none of which show any strong patterns. In Figure 4 we show the univariate coefficients of FEWS NET IPC, NDVI, rainfall, maximum temperature, diarrhoea admissions, and diesel price all compared to the univariate admissions coefficients for each subcounty; equivalent plots for the remaining variables are in supplementary Figure S20. The correlations between the coefficients are generally weak, with Pearson correlation coefficients for each of them at forecast horizons of 1 and 3 shown in supplementary Figures S21 and S22. For example the correlation between the coefficients across the subcounties between the admissions and NDVI for a forecast horizon of three months was 0.03 (95% CIs: −0.26 to 0.32), which shows no correlation. Similar results hold if we add in covariates (multi-variable regression), we do so both with time-lagged versions of the input variable of interest and with all the variables, with results (corresponding to Figures 3) shown in supplementary Figures S23 and S24.

**Fig. 4.**
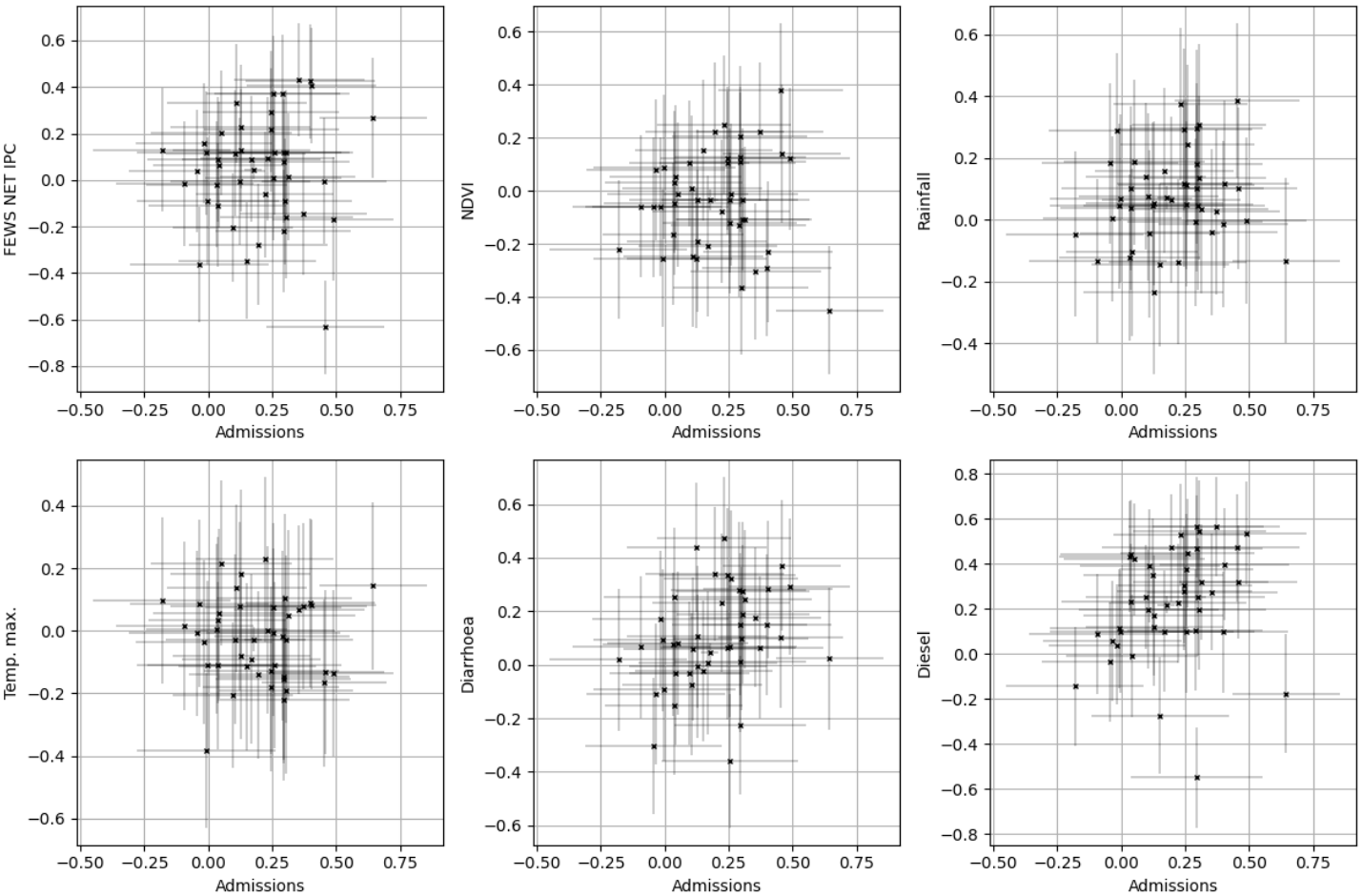
Univariate coefficient for each subcounty of FEWS NET IPC, NDVI, rainfall, maximum temperature, diarrhoea admissions, and diesel prices all compared to the univariate coefficients of the admissions. All coefficients are for a forecast horizon of three months.

While it is more difficult to assess the feature importance for XGBoost we show retained features for a three month forecast horizon in Figure 5, with an equivalent for a one month forecast horizon in supplementary Figure S25. Due to the nature of the XGBoost approach and the way that feature importance is assessed there is some stochasticity in these assessments; however the range of different features utilised remains high. There are also hints here of clustering effects by county, in the sense of groups of subcounties where the selected features are consistent - for example Narok county has quite considerable similarity in the selected features, as does Kajiado. There are substantial differences in the number of included features for counties and subcounties: for some, only a few features are retained while for others almost all features are.

**Fig. 5.**
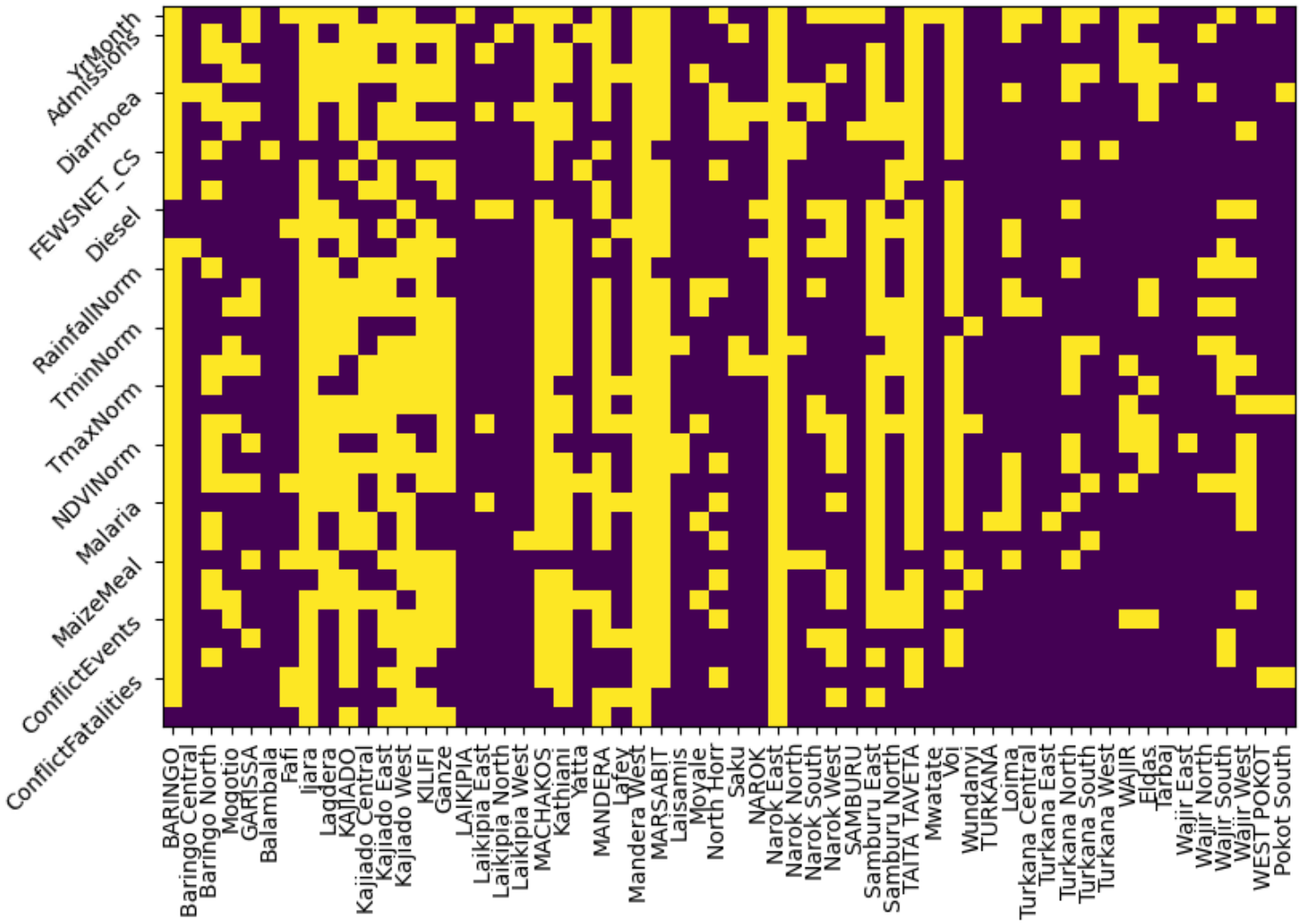
XGBoost feature importance at a forecast horizon of three months demonstrated with retained features (denoted as yellow in the plot) after applying feature selection through cross-validation. There are three squares associated with each variable for the time lags of 0, 1, 2 respectively. On the x-axis capitalised names are counties and those are trained with all the county data. The lower case names following each county name are the subcounties associated with the preceding county. For example BARINGO is a county and Baringo Central, Baringo North and Mogotio are all subcounties within Baringo county.

### 3.3 Temporal concept drift

To demonstrate the problem of temporal concept drift we show the univariate regression admissions coefficient of the three month forecast horizon calculated from a 30-month rolling time window (see Section 2.4 for details) for three example subcounties; the pattern of the autocorrelation for these subcounties varies substantially. For Balambala (left plot) the autocorrelation remains constant with a coefficient of around −0.2; for this subcounty, in this period, for this variable, temporal concept drift is not a problem. However, for both Baringo Central (middle plot) and Fafi (right plot) there is substantial change in the coefficients dependent on when the model was trained. For Baringo Central if training from 2017.5 up to around 2018.5 the coefficient remains relatively stable at approximately −0.3 but by the end of 2020, only 18 months apart, the coefficient has changed to around +0.4; therefore there is a potentially significant temporal concept drift. There is an inverted effect for Fafi, where in the earlier period the coefficient is slightly positive (around 0.15) and in the later period becomes negative (around −0.5). For both Fafi and Baringo Central the univariate coefficient depends substantially on the time-period of data that is available to train on.

From the univariate temporal drift analysis we extracted the maximum and minimum coefficients across the time-period; in Figure 7 we plotted these for admissions, FEWS NET IPC and NDVI for all subcounties and a forecast horizon of three months, uncertainties on coefficients are included for a small number of subcounties to prevent the figures becoming difficult to read. There are large uncertainties in the coefficients but considerable temporal concept drift is still evident with substantial changes between maximum and minimum coefficients with time. For example, for one sub-county (Laikipia West) the coefficient on the IPC scores ranges from 0.62 (0.31 to 0.92) to −0.47 (−0.80 to −0.13); or from there being a moderate positive correlation to a moderate negative correlation. In supplementary Figure S26 we show an equivalent plot for a time-lag of one month, and similar substantial changes in minimum to maximum coefficients can be observed.

In Figure 8 we show maximum versus minimum coefficients for all of the variables for three of the four subcounties of Marsabit county with a time-lag of three months.

Equivalent plots for a time-lag of 1 month are shown in supplementary Figure S27. Given these subcounties are from the same county there are quite substantial differences in the patterns between maximum and minimum coefficients which demonstrates the combinational challenge of spatial and temporal concept drift.

### 3.4 Model predictions

We summarise the overall forecasting results for a forecast horizon of 3 months in Table 1, these are the overall RMSE when concatenating over the test sets of all the subcounties; equivalent results for a forecast horizon of 1 month are in supplementary Table S2.

**Table 1.**
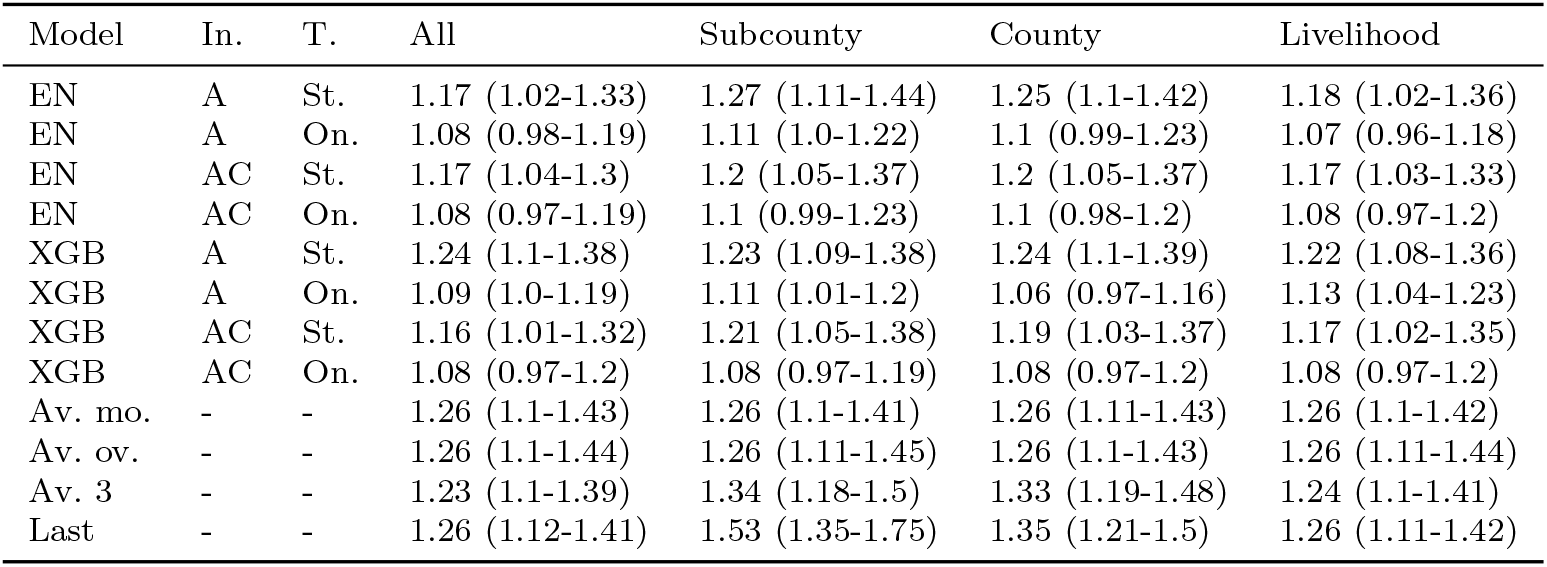
Forecasting results (RMSE) for a forecast horizon of 3 months for the test sets concatenated over all the subcounties, uncertainties at 95% are shown in the brackets. Results for the elastic net (EN) and XGBoost (XGB) are shown as are baselines for average per month (Av. mon.), averaged overall (Av. ov.), the average of the last three months (Av. 3), and the last value (Last). For the models results are shown for different inputs (labelled as In.) with all (A) and autocorrelation alone (AC) shown. Also for the models the training approach using either standard training (St.) or ongoing (On.) is also shown.

| Model | In. | T. | All | Subcounty | County | Livelihood |
|---|---|---|---|---|---|---|
| EN | A | St. | 1.17 (1.02-1.33) | 1.27 (1.11-1.44) | 1.25 (1.1-1.42) | 1.18 (1.02-1.36) |
| EN | A | On. | 1.08 (0.98-1.19) | 1.11 (1.0-1.22) | 1.1 (0.99-1.23) | 1.07 (0.96-1.18) |
| EN | AC | St. | 1.17 (1.04-1.3) | 1.2 (1.05-1.37) | 1.2 (1.05-1.37) | 1.17 (1.03-1.33) |
| EN | AC | On. | 1.08 (0.97-1.19) | 1.1 (0.99-1.23) | 1.1 (0.98-1.2) | 1.08 (0.97-1.2) |
| XGB | A | St. | 1.24 (1.1-1.38) | 1.23 (1.09-1.38) | 1.24 (1.1-1.39) | 1.22 (1.08-1.36) |
| XGB | A | On. | 1.09 (1.0-1.19) | 1.11 (1.01-1.2) | 1.06 (0.97-1.16) | 1.13 (1.04-1.23) |
| XGB | AC | St. | 1.16 (1.01-1.32) | 1.21 (1.05-1.38) | 1.19 (1.03-1.37) | 1.17 (1.02-1.35) |
| XGB | AC | On. | 1.08 (0.97-1.2) | 1.08 (0.97-1.19) | 1.08 (0.97-1.2) | 1.08 (0.97-1.2) |
| Av. mo. | - | - | 1.26 (1.1-1.43) | 1.26 (1.1-1.41) | 1.26 (1.11-1.43) | 1.26 (1.1-1.42) |
| Av. ov. | - | - | 1.26 (1.1-1.44) | 1.26 (1.11-1.45) | 1.26 (1.1-1.43) | 1.26 (1.11-1.44) |
| Av. 3 | - | - | 1.23 (1.1-1.39) | 1.34 (1.18-1.5) | 1.33 (1.19-1.48) | 1.24 (1.1-1.41) |
| Last | - | - | 1.26 (1.12-1.41) | 1.53 (1.35-1.75) | 1.35 (1.21-1.5) | 1.26 (1.11-1.42) |

Model performance is insufficiently high to enable usable forecasts to be made. Our models significantly outperform any of the baselines we assessed at the aggregated level, with the best performing baseline giving an overall RMSE of 1.23 (1.10-1.39) compared to 1.06 (0.97-1.16) of the best performing model. In supplementary Figure S28 we show a comparison between the RMSE per subcounty for the forecasting models and baselines, the models provide modestly better performance than the baselines for most of the subcounties.

There is no significant difference in performance between XGBoost and the elastic net regression with RMSEs for the best performing versions of 1.06 (0.97-1.16) and 1.07 (0.96-1.18) respectively. This general trend of similarity of performance holds consistently across the different versions of the models with similar performance. We also see similar performance between XGBoost and elastic net regression for the individual subcounties (supplementary Figure S29).

We attempted to compensate for temporal concept drift by performing ongoing training which generally produced a modest improvement in performance, for example with XGBoost using all inputs and no additional spatial information the RMSE for standard training and ongoing training were 1.24 (1.10-1.38) and 1.09 (1.00-1.19) respectively. A similar improvement was apparent for elastic net. We show the RMSEs for individual subcounties for ongoing compared to standard training in supplementary Figure S30, where RMSEs for most subcounties are similar with a few subcounties showing larger differences.

There was little difference in performance when including exogenous variables, with models trained using autocorrelation alone producing similar performance as those with all inputs. For example, the XGBoost modelled trained with no additional spatial information, and in an ingoing manner, produced RMSEs for all inputs and on just autocorrelation of 1.09 (1.01-1.19) and 1.08 (0.97-0.20) respectively. Plots for the RMSE for individual subcounties showing the differences in performance for using just autocorrelation or using all the inputs are in supplementary Figure S31.

There were no significant differences for XGBoost when including spatial information regardless of whether all inputs were included or whether standard or ongoing training was utilised. For the elastic net regression models there were also not significant differences in performance for different spatial approaches, it is important to note that the elastic net was trained separately for the different spatial scales so there was much less data especially for the subcounty version.

## 4 Discussion

This study examined whether spatial and temporal concept drift may help explain why ML models struggle to forecast malnutrition admissions and why adding exogenous predictors often provides limited benefit. Across univariate, multivariable and XGBoost analyses, relationships between predictors and admissions varied substantially between locations and over time. These results do not establish concept drift as the sole cause of poor forecasting performance, but they show that the conventional practice of geographically pooled observations sharing a common effect from a predictor–outcome relationship is often questionable in this setting.

The spatial results expose a fundamental trade-off between data quantity and data relevance. Training models separately for individual subcounties limits spatial drift but leaves relatively little data, with correspondingly uncertain coefficient estimates and generally weaker forecasting performance. Pooling subcounties increases sample size, but the coefficients and selected features vary considerably across locations (Figures 3–5). Importantly, these differences do not form simple or consistent patterns: a subcounty with a strong lagged-admissions relationship does not necessarily show correspondingly strong relationships for IPC, NDVI or other predictors. This makes it difficult to identify groups for which a single set of relationships can be assumed.

Temporal drift creates an analogous problem. In several subcounties, both the magnitude and sign of associations changed materially depending on the training period (Figures 6–8). For example, the lagged-admissions coefficient for Baringo Central changed from negative to positive over a relatively short period. Temporal drift was also heterogeneous across locations and predictors: some subcounties showed comparatively stable relationships while others changed substantially. Extending the training window backwards can therefore add observations while simultaneously adding relationships that are less representative of the forecast period.

**Fig. 6.**
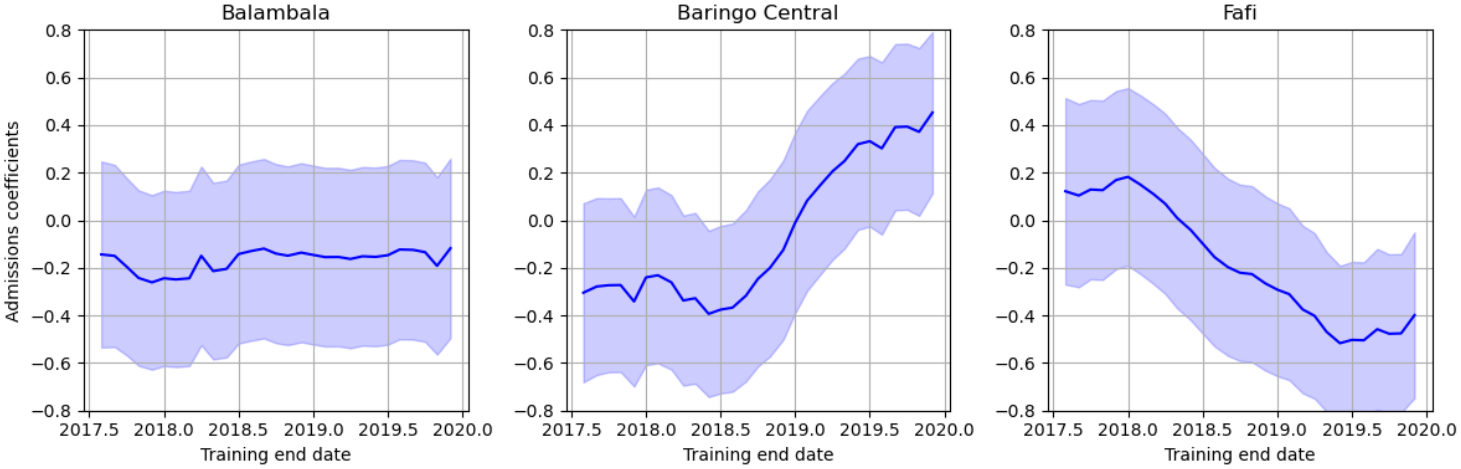
Temporal concept drift for a forecast horizon of three months for the admissions coefficient for three example subcounties. The univariate relationship is found for a 30 month sliding window with the last date given as the x-axis value. For Balambala (left) the coefficient does not change with time, staying at around a constant −0.2. For Baringo Central there is a significant change in coefficient values at a training end date of around 2018.5 where the coefficient increases significantly. Conversely for Fafi the coefficient drops substantially after around 2018.

**Fig. 7.**
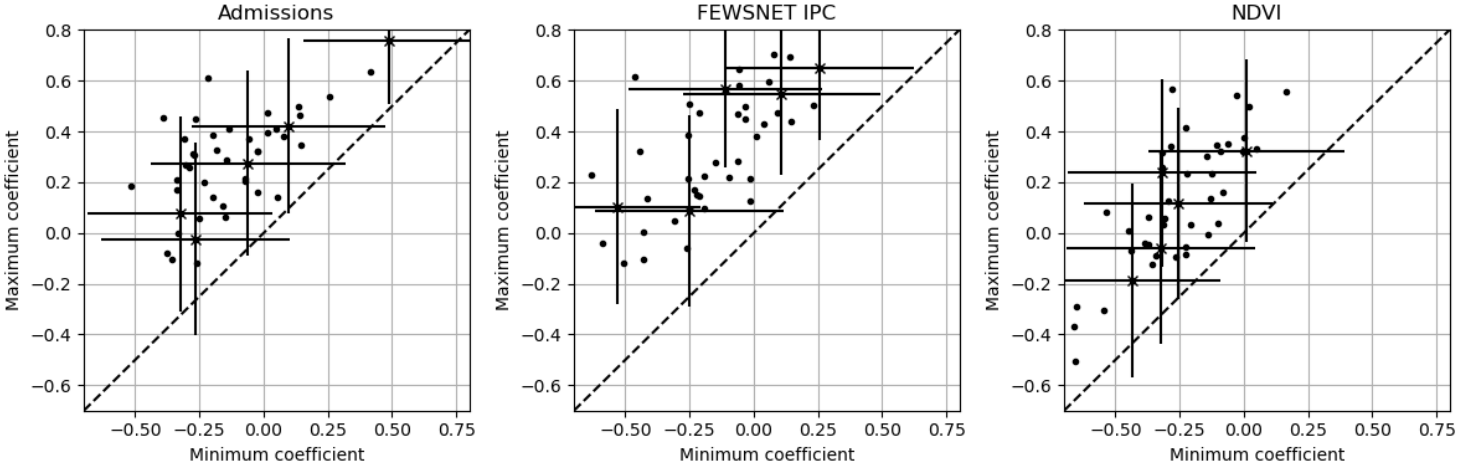
The maximum versus minimum coefficients for three variables (admissions, FEWSNET IPC, NDVI) for all subcounties at a forecast horizon of three months when using a 30-month sliding window to train the univariate regression.

**Fig. 8.**
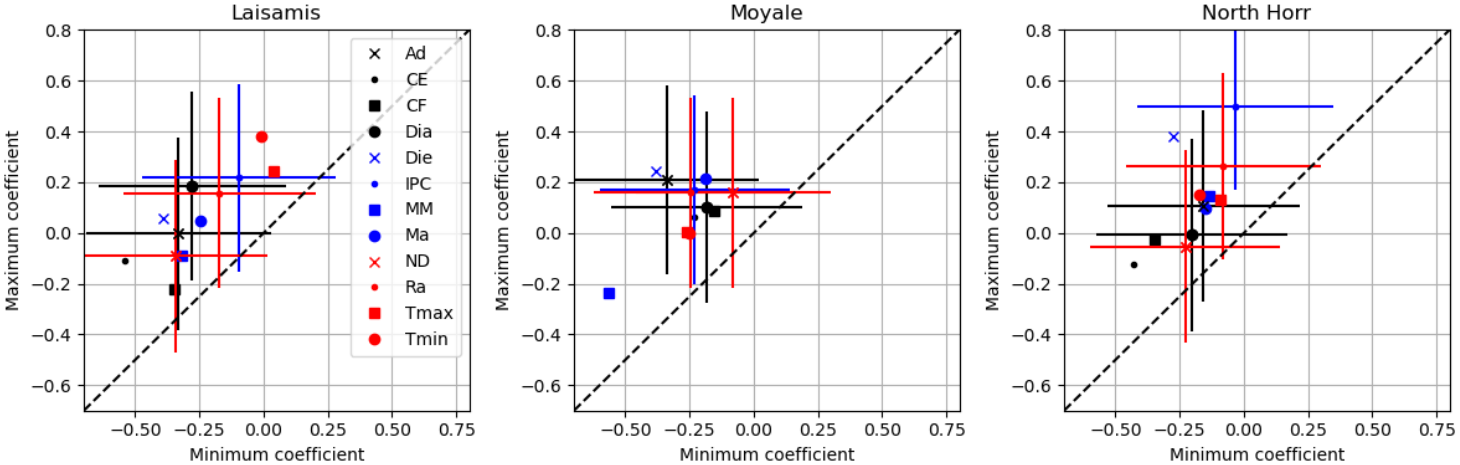
Maximum versus minimum coefficients for all variables at a forecast horizon of three month for an example three subcounties from the county of Marsabit.

These findings provide a possible explanation for the dominance of autocorrelation reported in previous malnutrition forecasting studies [5, 12, 13]. If associations between exogenous variables and admissions differ across locations or time, a pooled model has limited ability to assign a single stable importance to those variables. Lagged admissions may consequently remain the most consistently useful predictor. This interpretation is supported by our forecasting experiments, where adding exogenous variables produced little improvement over using lagged admissions alone.

We also considered whether a flexible non-linear model could implicitly accommodate this heterogeneity. XGBoost did not consistently outperform elastic net, even when spatial information was provided. This does not imply that non-linearities or interactions are absent; rather, the available data may be insufficient for a flexible model to learn relationships that generalise across heterogeneous locations and periods. Greater model complexity alone therefore appears unlikely to solve the problem.

The mitigation experiments produced mixed results. Ongoing training gave consistent but modest improvements, supporting the idea that incorporating the most contemporary observations can reduce the effect of temporal drift. By contrast, grouping subcounties by county or livelihood zone did not consistently improve performance. These groupings may be too coarse to capture the dimensions along which predictor–admission relationships are actually similar. More targeted, data-driven or domain-informed approaches for identifying comparable locations may be needed.

The broader implication is not that additional data are unhelpful. Indeed, models trained on pooled data often performed better than models trained on individual sub-counties. Rather, the results suggest that *how* additional data are combined matters when relationships are non-stationary. Future work could explore more appropriate spatial groupings, stronger feature selection and predictors or composite indicators that are more stable across settings. Some apparent drift may also arise from omitted variables. For example, the effect of rainfall may depend on irrigation, livelihoods or access to markets, all of which can vary spatially and temporally. Or that admissions can be related to the implementation of outreach programmes referring people to health facilities. Incorporating domain knowledge and causal structure may therefore help identify missing modifiers and improve transferability.

Another area for future work would be to investigate the effect of smaller spatial scale for the admissions data, we used data aggregated at the subcounty level; it would be interesting to explore data at a higher spatial level. Also we analysed patterns from combined MAM and SAM, it would be plausible that different patterns may emerge for MAM and SAM separately. We did perform a univariate analysis of SAM, finding the same type of patterns as the combined data, but a more in-depth analysis could be valuable.

Several limitations should be considered. The analysis covers 44 subcounties in Kenya and may not generalise to regions with different levels of heterogeneity or to other measures of malnutrition. Feature importance, particularly for XGBoost, is an imperfect measure of concept drift, and poor forecasting may also reflect noisy admissions data, missing predictors or limited sample size. We therefore interpret concept drift as a plausible and empirically supported contributor rather than a definitive explanation. Nevertheless, the consistency of the spatial and temporal evidence, together with the modest benefit of ongoing training, indicates that concept drift deserves explicit consideration in future malnutrition forecasting systems.

## 5 Conclusions

Forecasting malnutrition admissions to an actionable level remains difficult, and previous studies similarly report limited gains from exogenous variables beyond time-lagged measures of malnutrition. Our results provide evidence that spatial and temporal concept drift are plausible contributors to this difficulty: relationships between predictors and admissions vary across both locations and time, reducing the extent to which additional historical or geographically pooled data can be assumed to represent the target setting.

The mitigation approaches explored here produced only modest improvements. Ongoing training helped to reduce the effect of temporal drift, whereas grouping data by county or livelihood zone did not consistently overcome spatial drift. These findings do not imply that additional data are unhelpful; rather, they show that how data are combined matters when the underlying predictor–outcome relationships are non-stationary.

The main implication is that forecasting approaches for malnutrition admissions should explicitly account for spatial and temporal heterogeneity. Further gains may require better ways of identifying comparable regions, selecting or constructing more stable predictors, and incorporating domain knowledge or causal structure. Concept drift therefore provides both a potential explanation for the limited performance of current models and a practical direction for future forecasting research.

## Data Availability

The malnutrition, malaria, diarrhoea admissions data requires access to the Kenya Health Information System which needs to be requested by interested researchers. All other data is available online with web-links provided in the manuscript.

https://fews.net/

https://www.chc.ucsb.edu/data/chirps3

https://www.chc.ucsb.edu/data/chirts-era5

https://www.earthdata.nasa.gov/topics/land-surface/normalized-difference-vegetation-index-ndvi

https://acleddata.com/

## Acknowledgements

This work was conducted as part of the Health System Resilience to Climate-related Surges (HERCS) project.

HERCS is part of the Climate Adaptation and Resilience (CLARE) programme, funded mostly (about 90%) by UK Aid through the Foreign Commonwealth and Development Office (FCDO), and co-funded by the International Development Research Centre (IDRC), Canada.

This work was funded by UK aid from the UK government and by the International Development Research Centre (IDRC), Ottawa, Canada, as part of the Climate Adaptation and Resilience (CLARE) research programme. The views expressed herein do not necessarily represent those of the UK government, IDRC or its Board of Governors.

