## Supplemental material for "Analysing Spatial and Temporal Concept Drift in Forecasting Malnutrition Admissions in Kenya"

### 1 Supplementary methodology

#### 1.1 Data and pre-processing

We select potential features from a review of the literature on the relationship to malnutrition together with availability of data. We used: time-lagged values of malnutrition admissions, FEWSNET IPC current situation scores, diarrhoea admissions, malaria admissions, diesel prices, maize meal prices, normalised difference vegetation index (NDVI), maximum temperatures, minimum temperatures, total rainfall, number of conflict events, and number of conflict fatalities.

All inputs and output data was standardised to produce z-scores. This was done by subcounty, using the training set to calculate the mean and standard deviation, which was subsequently applied to the testing set to standardise with reference to the training set. For the NDVI, temperatures and total rainfall the standardisation was performed by month such that they become the standard deviations away from the mean for that month and that subcounty. For the other variables standardisation was performed over the whole time-period of the training set.

Some of the data was provided at sub-county level: malnutrition admissions, malaria admissions, diarrhoea admissions while diesel and maize meal prices are both provided at country-level, for these no processing is done as they can be directly used at subcounty level. Rainfall, temperature and NDVI data was accessed at monthly level so no time-related processing was required; for the spatial scale these variables are provided at high spatial scale so we averaged them over the area of the specified subcounty. Conflict events and fatalities are provided at the monthly level and are available per subcounty, we summed all of each by month and by subcounty.

For the univariate analysis we use time lags between the input variable and malnutrition admissions of 1, 2, 3, 4, 5, 6 months. For the covariate analysis and for the forecasting models we target the malnutrition admissions at 1, 3, 6 months into the future using the inputs at time lags (number of months before the present) of 0, 1, 2.

Table S1: The features used in statistical analysis and training the model.

| Predictor | Source |
|---|---|
| Malnutrition admissions | Kenya Health Information System - need to request access |
| Malaria admissions | Kenya Health Information System - need to request access |
| Diarrhoea admissions | Kenya Health Information System - need to request access |
| FEWSNET IPC | `https://fews.net/` |
| Diesel price | `https://fews.net/` |
| Maize meal price | `https://fews.net/` |
| Total rainfall | `https://www.chc.ucsb.edu/data/chirps3` |
| Maximum temperature | `https://www.chc.ucsb.edu/data/chirts-era5` |
| Minimum temperature | `https://www.chc.ucsb.edu/data/chirts-era5` |
| NDVI | `https://www.earthdata.nasa.gov/topics/land-surface/normalized-difference-vegetation-index-ndvi` |
| Conflict events | `https://acleddata.com/` |
| Conflict fatalities | `https://acleddata.com/` |

For the spatial data relating to location of Kenya's boundaries including counties and subcounties we used data from `https://data.humdata.org/dataset/`. We define livelihood zones via the FEW NET (`https://fews.net/`) classifications downloaded as .shp files; we map the livelihood zone designations to the subcounties and we define each subcounty using the livelihood zone which makes up the majority (or the largest fraction) the subcounty. We use three overall livelihood zones taken from the FEW NET downloaded data: PS; combined AG and AN; and SH, in Figure S1.

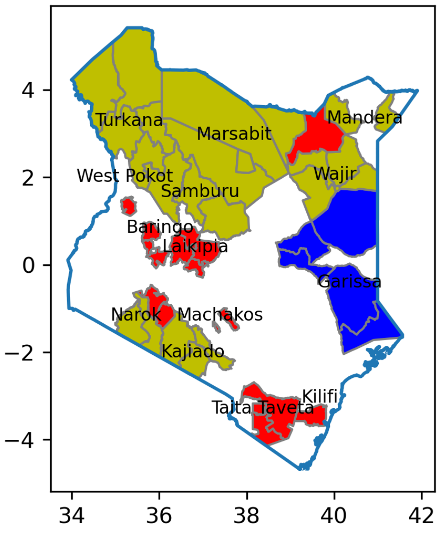


Figure S1: Defined livelihood zones as defined by FEWS NET classes: PS, AG/AN, and SH.

### 2 Supplementary results

#### 2.1 Admissions data

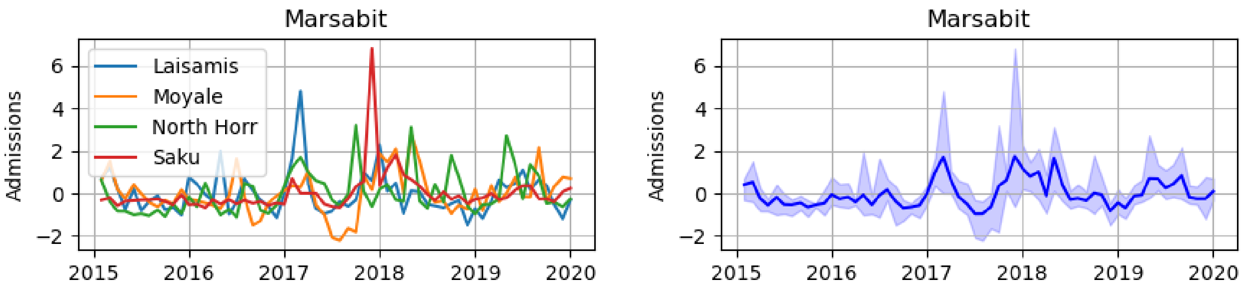


Figure S2: Example standardised (z-scores) admissions data from the county Marsabit. Left) the number of malnutrition admissions per month for the four subcounties making up Marsabit county (Laisamis, Moyale, North Horr, and Saku). Right) The mean (blue line) value of admissions of the subcounties per month with the range of the admissions shown as blue shading.

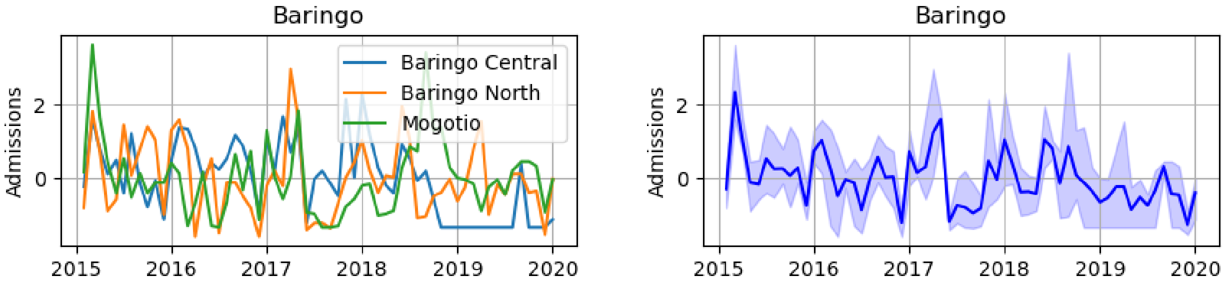


Figure S3: Example standardised (z-scores) admissions data from the county Baringo. Left) the number of malnutrition admissions per month for the three subcounties making up Baringo county. Right) The mean (blue line) value of admissions of the subcounties per month with the range of the admissions shown as blue shading.

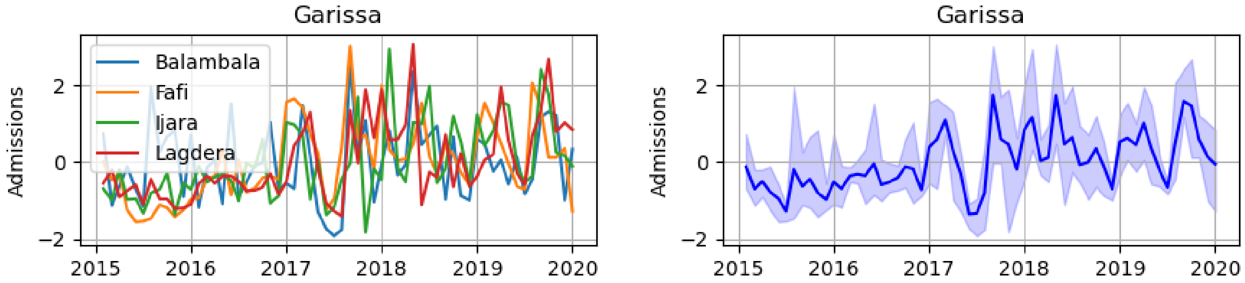


Figure S4: Example standardised (z-scores) admissions data from the county Garissa. Left) the number of malnutrition admissions per month for the four subcounties making up Garissa county. Right) The mean (blue line) value of admissions of the subcounties per month with the range of the admissions shown as blue shading.

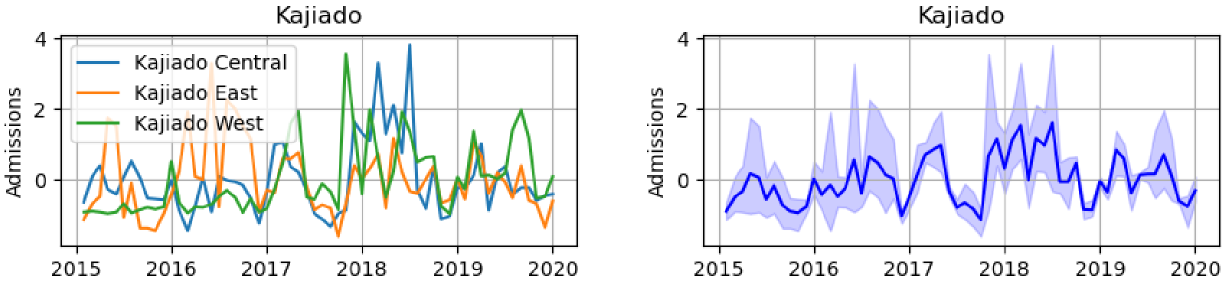


Figure S5: Example standardised (z-scores) admissions data from the county Kajiado. Left) the number of malnutrition admissions per month for the four subcounties making up Kajiado county. Right) The mean (blue line) value of admissions of the subcounties per month with the range of the admissions shown as blue shading.

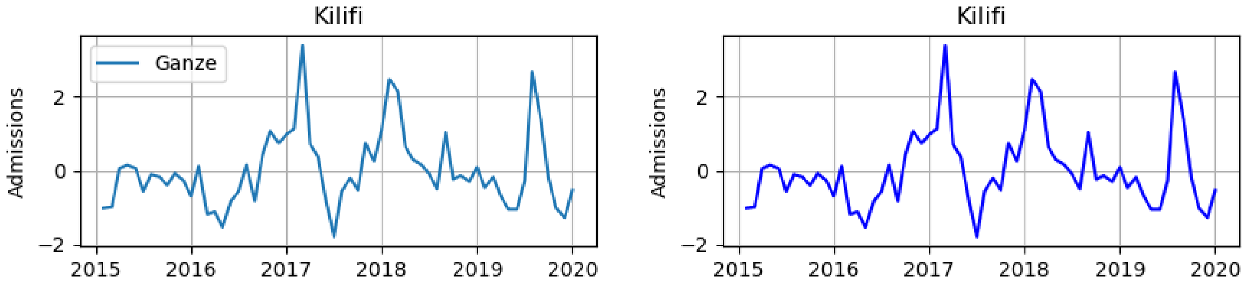


Figure S6: Example standardised (z-scores) admissions data from the county Kilifi. Left) the number of malnutrition admissions per month for the one subcounty making up Kilifi county. Right) The mean (blue line) value of admissions of the subcounty.

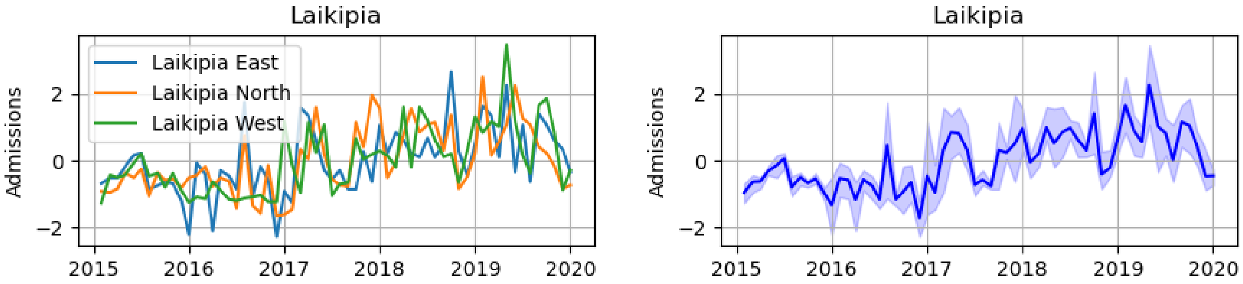


Figure S7: Example standardised (z-scores) admissions data from the county Laikipia. Left) the number of malnutrition admissions per month for the three subcounties making up Laikipia county. Right) The mean (blue line) value of admissions of the subcounties per month with the range of the admissions shown as blue shading.

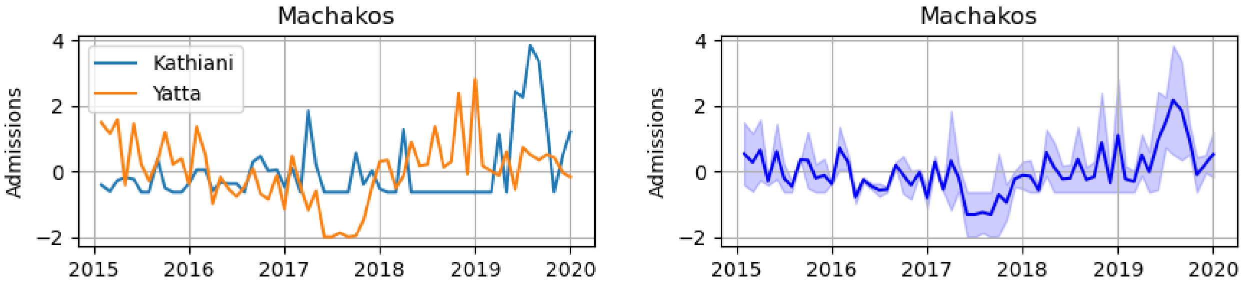


Figure S8: Example standardised (z-scores) admissions data from the county Machakos. Left) the number of malnutrition admissions per month for the two subcounties making up Machakos county. Right) The mean (blue line) value of admissions of the subcounties per month with the range of the admissions shown as blue shading.

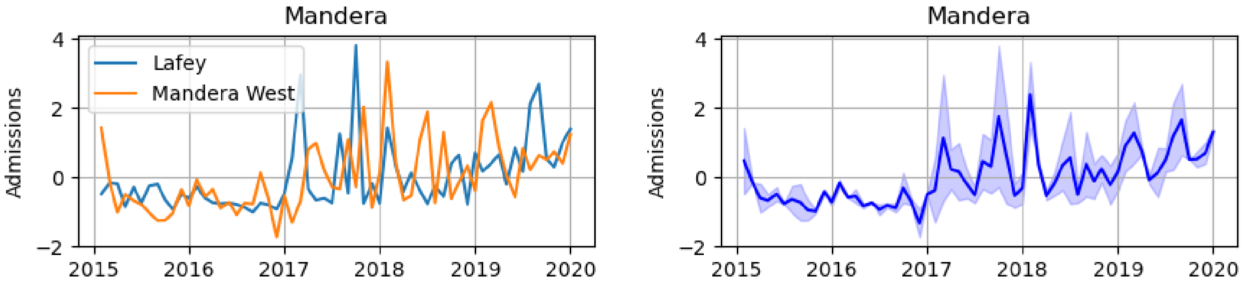


Figure S9: Example standardised (z-scores) admissions data from the county Mandera. Left) the number of malnutrition admissions per month for the two subcounties making up Mandera county. Right) The mean (blue line) value of admissions of the subcounties per month with the range of the admissions shown as blue shading.

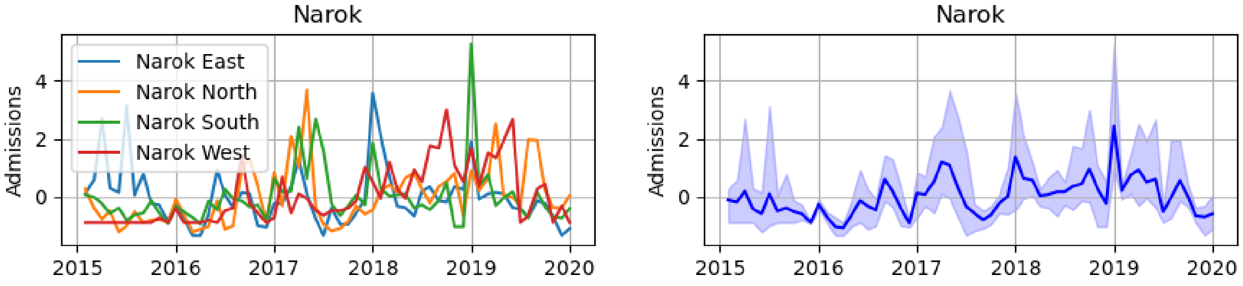


Figure S10: Example standardised (z-scores) admissions data from the county Narok. Left) the number of malnutrition admissions per month for the four subcounties making up Narok county. Right) The mean (blue line) value of admissions of the subcounties per month with the range of the admissions shown as blue shading.

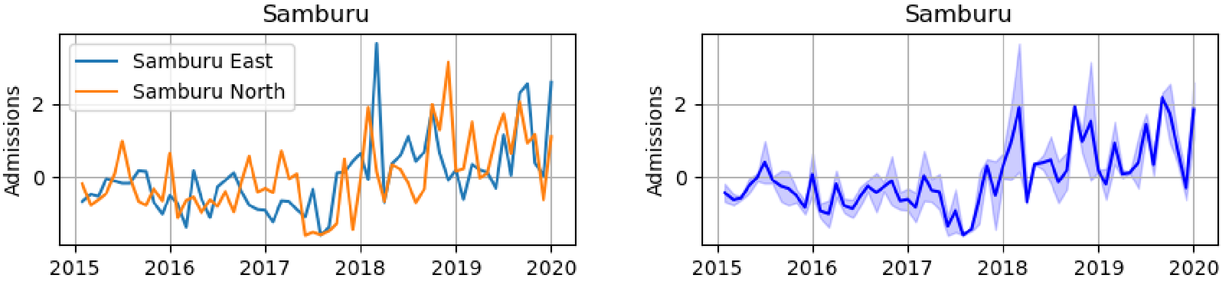


Figure S11: Example standardised (z-scores) admissions data from the county Samburu. Left) the number of malnutrition admissions per month for the two subcounties making up Samburu county. Right) The mean (blue line) value of admissions of the subcounties per month with the range of the admissions shown as blue shading.

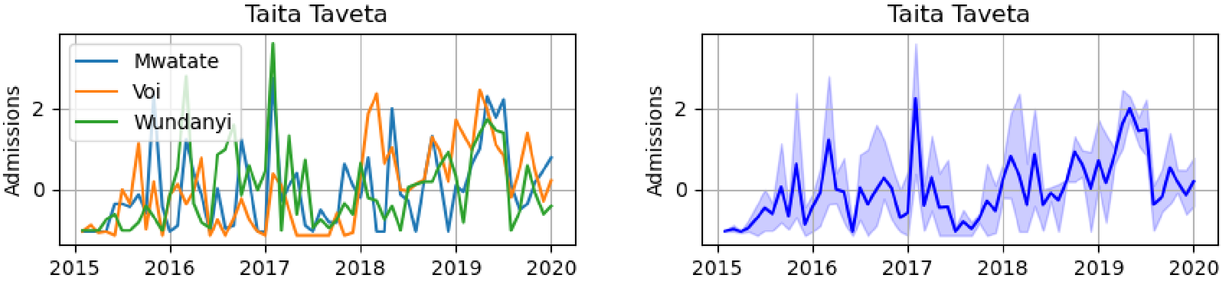


Figure S12: Example standardised (z-scores) admissions data from the county Taita Taveta. Left) the number of malnutrition admissions per month for the three subcounties making up Taita Taveta county. Right) The mean (blue line) value of admissions of the subcounties per month with the range of the admissions shown as blue shading.

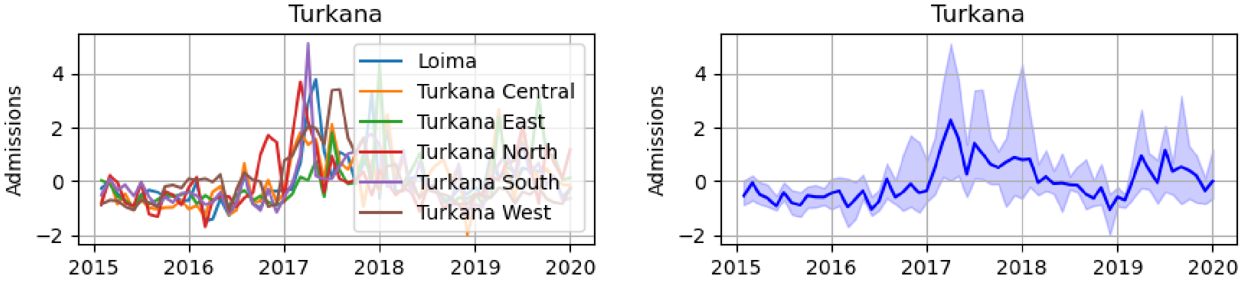


Figure S13: Example standardised (z-scores) admissions data from the county Turkana. Left) the number of malnutrition admissions per month for the six subcounties making up Turkana county. Right) The mean (blue line) value of admissions of the subcounties per month with the range of the admissions shown as blue shading.

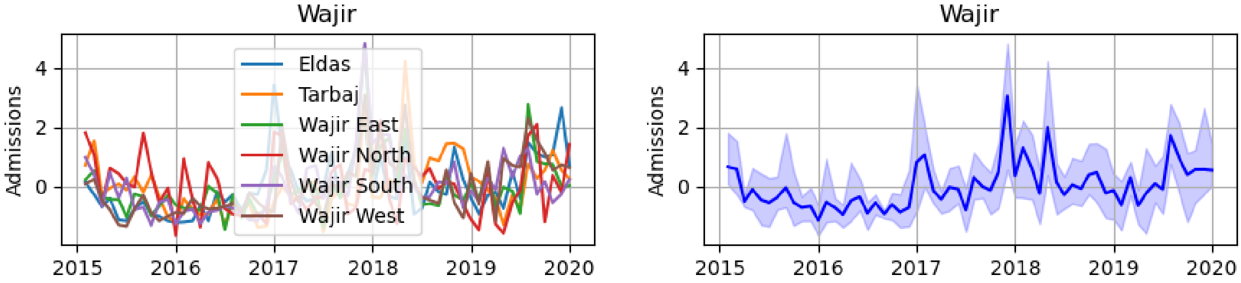


Figure S14: Example standardised (z-scores) admissions data from the county Wajir. Left) the number of malnutrition admissions per month for the six subcounties making up Wajir county. Right) The mean (blue line) value of admissions of the subcounties per month with the range of the admissions shown as blue shading.

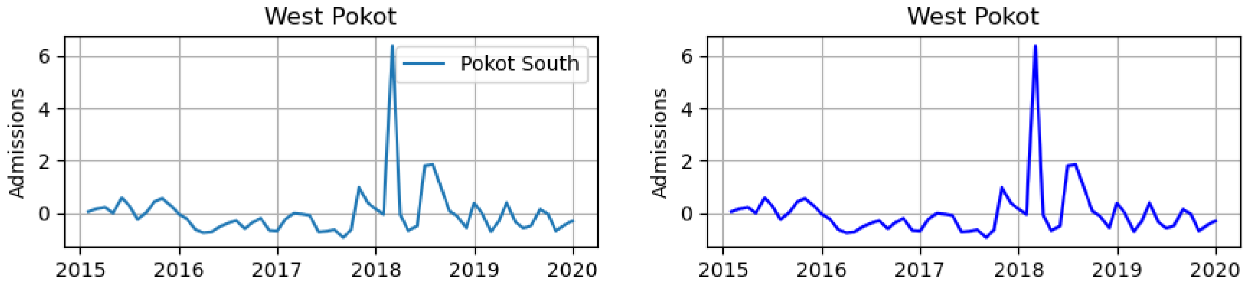


Figure S15: Example standardised (z-scores) admissions data from the county West Pokot. Left) the number of malnutrition admissions per month for the subcounty making up West Pokot county. Right) The mean (blue line) value of admissions of the subcounty.

Figure S16 shows the Pearson correlation coefficients betwen all 45 subcounties included in the analysis. The subcounties are ordered by county with the county labelled at the top of the plot and the black lines showing the groups of counties. Some counties, such as Turkana, tend to show stronger correlations in admissions than other counties, such as Marsabit. There is a general pattern of greater correlation between subcounties from the same county than to subcounties outside the county.

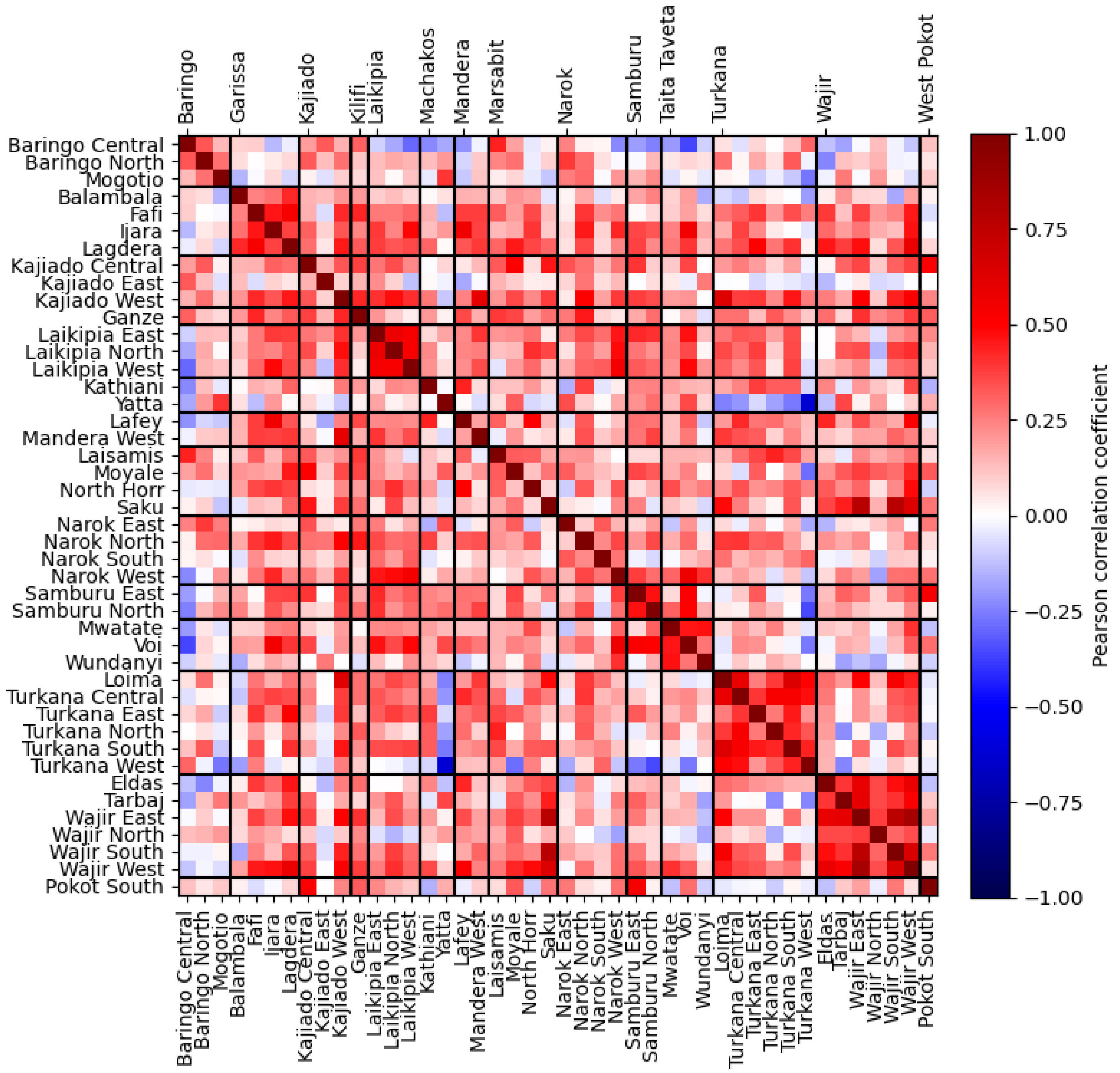


Figure S16: Pearson correlation coefficients between subcounties. Subcounties grouped together by counties labelled on the top of the plot.

### 2.2 Spatial concept drift

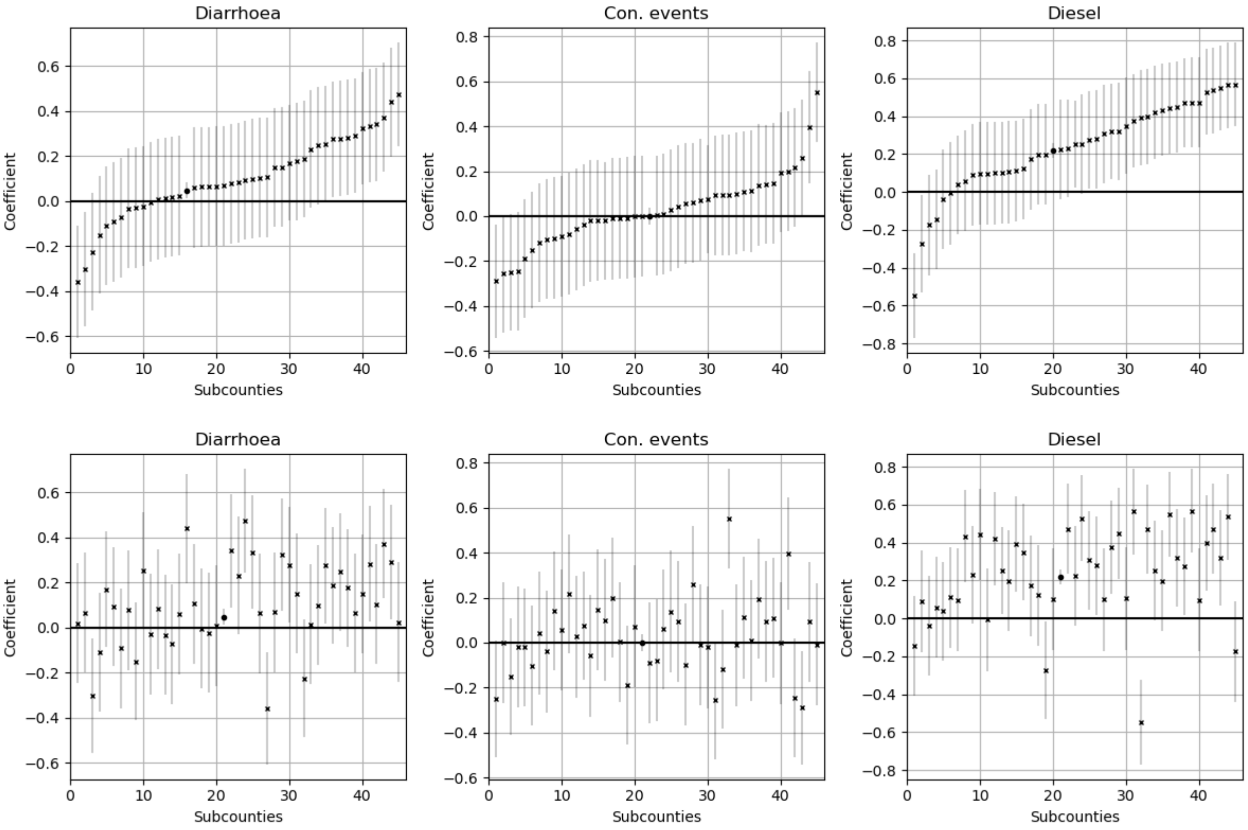


Figure S17: Univariate coefficients by subcounty for a forecast horizon of three months. Also shown is the coefficient if all data across the subcounties is concatenated, shown as a large circle marker and with much smaller confidence intervals. Top) The coefficients have been sorted separately for each variable to show the overall spread of the coefficients across the subcounties. Bottom) The coefficients have been sorted by the same ordering as for the admissions coefficients, to show the lack of obvious relationship if we order by the admissions coefficient.

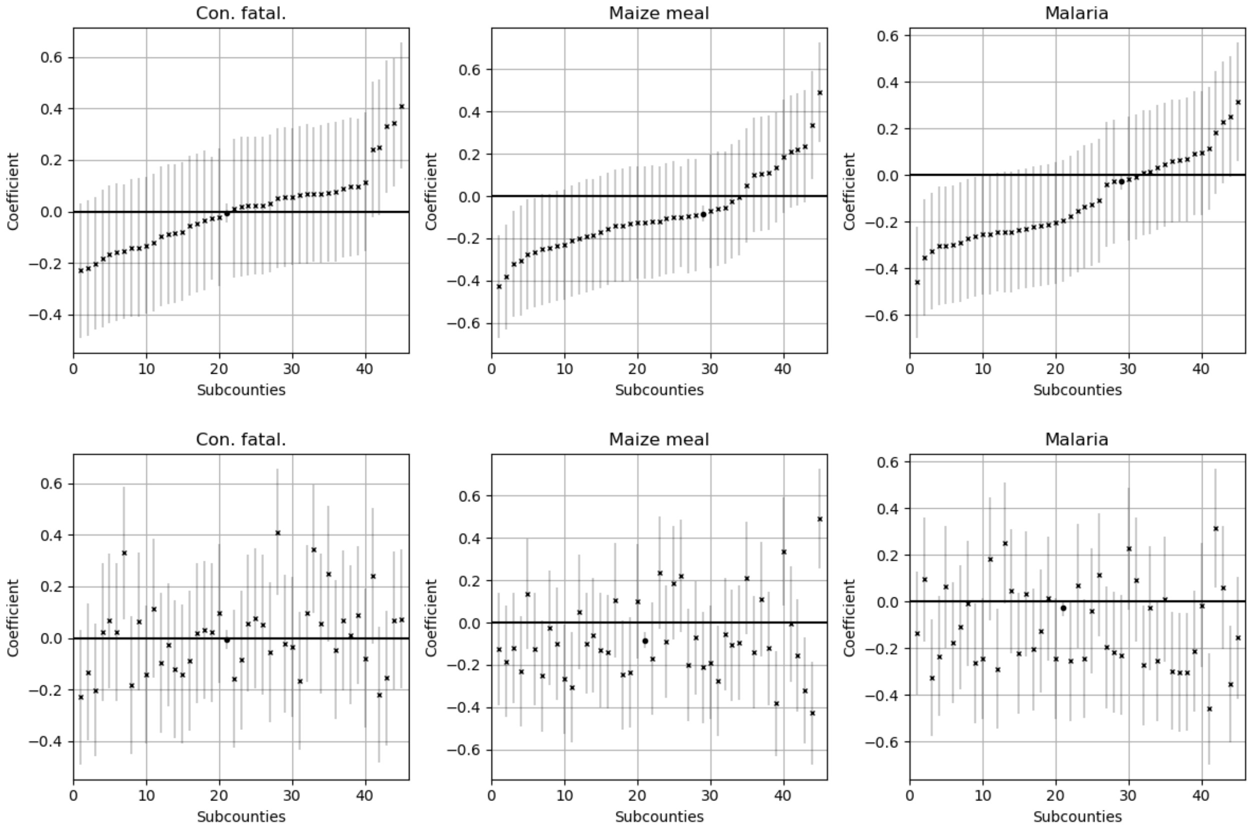


Figure S18: Univariate coefficients by subcounty for a forecast horizon of three months. Also shown is the coefficient if all data across the subcounties is concatenated, shown as a large circle marker and with much smaller confidence intervals. Top) The coefficients have been sorted separately for each variable to show the overall spread of the coefficients across the subcounties. Bottom) The coefficients have been sorted by the same ordering as for the admissions coefficients, to show the lack of obvious relationship if we order by the admissions coefficient.

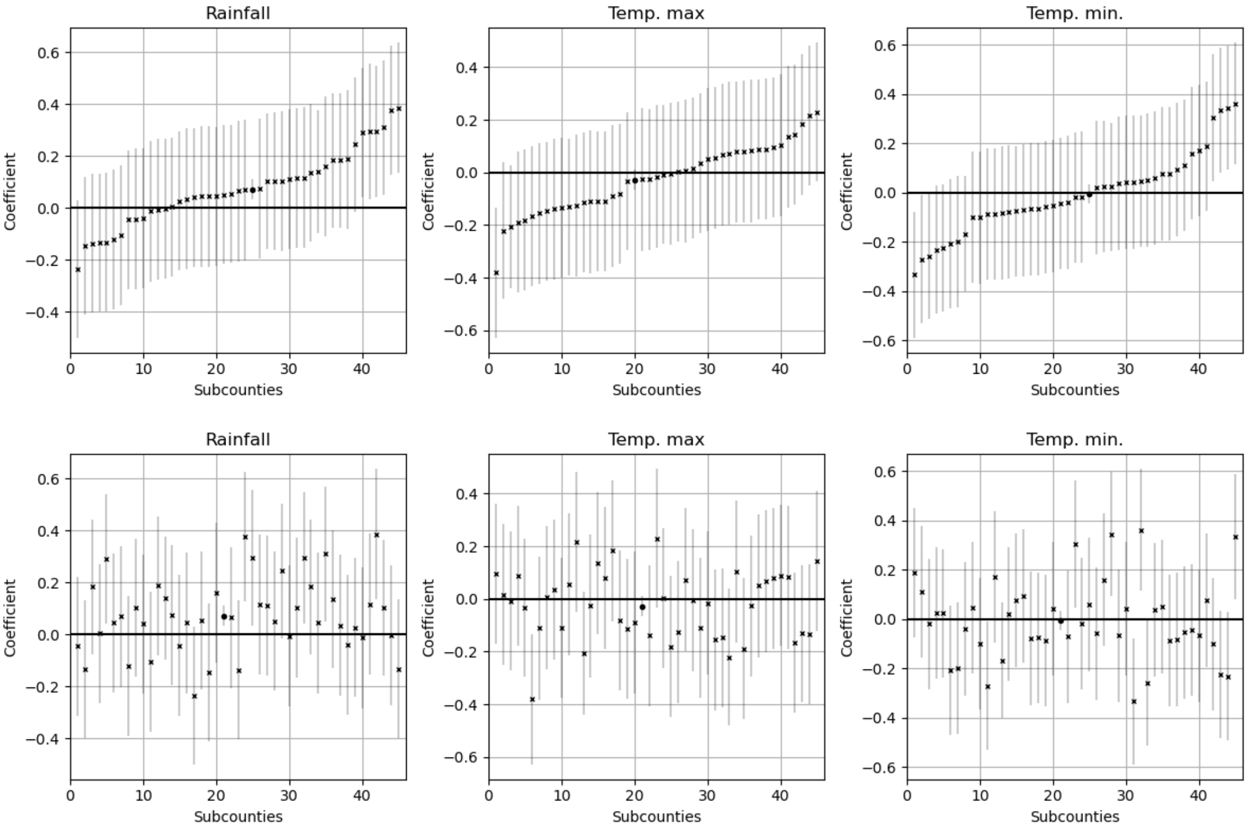


Figure S19: Univariate coefficients by subcounty for a forecast horizon of three months. Also shown is the coefficient if all data across the subcounties is concatenated, shown as a large circle marker and with much smaller confidence intervals. Top) The coefficients have been sorted separately for each variable to show the overall spread of the coefficients across the subcounties. Bottom) The coefficients have been sorted by the same ordering as for the admissions coefficients, to show the lack of obvious relationship if we order by the admissions coefficient.

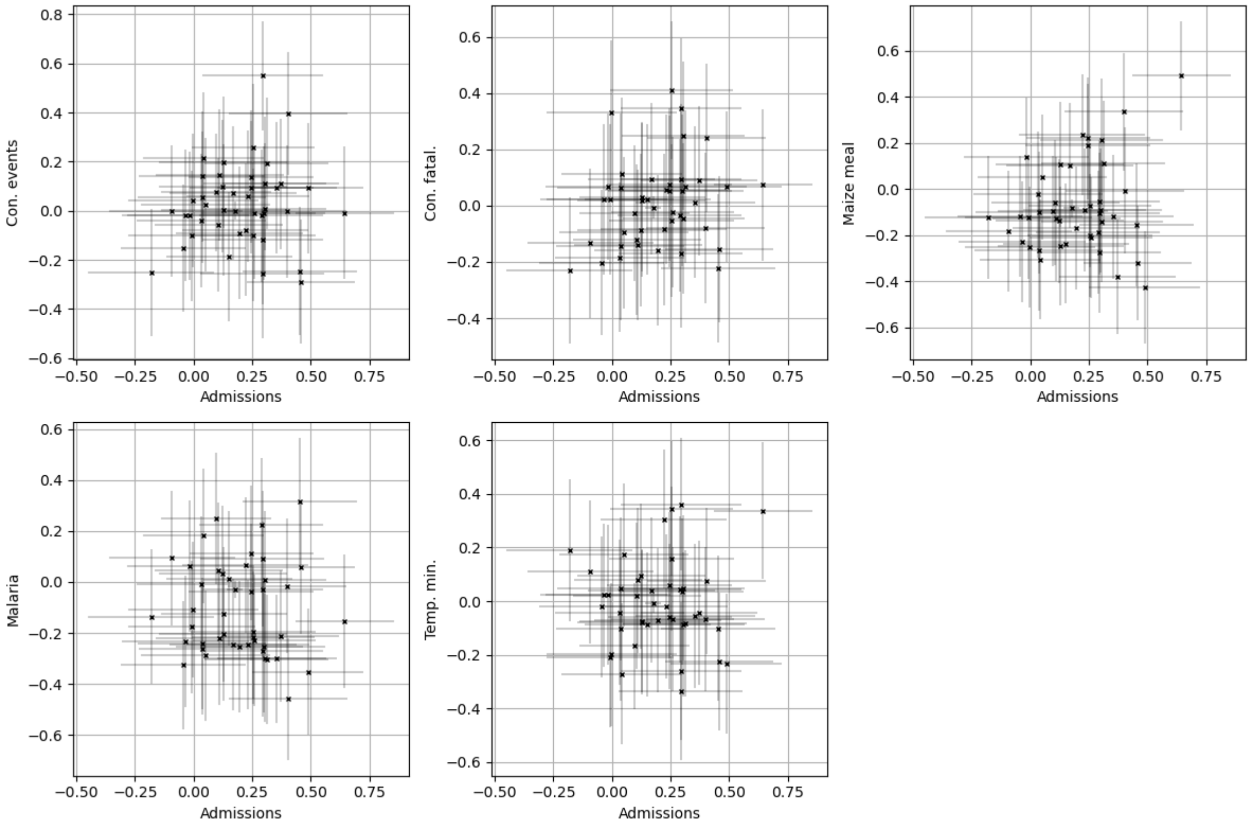


Figure S20: Coefficients of number of conflict events (Con. events), number of conflict fatalities (Con. fatal.), maize meal prices, malaria admissions and minimum temperature all compared to the coefficients of the admission. All coefficients are for a forecast horizon of three months.

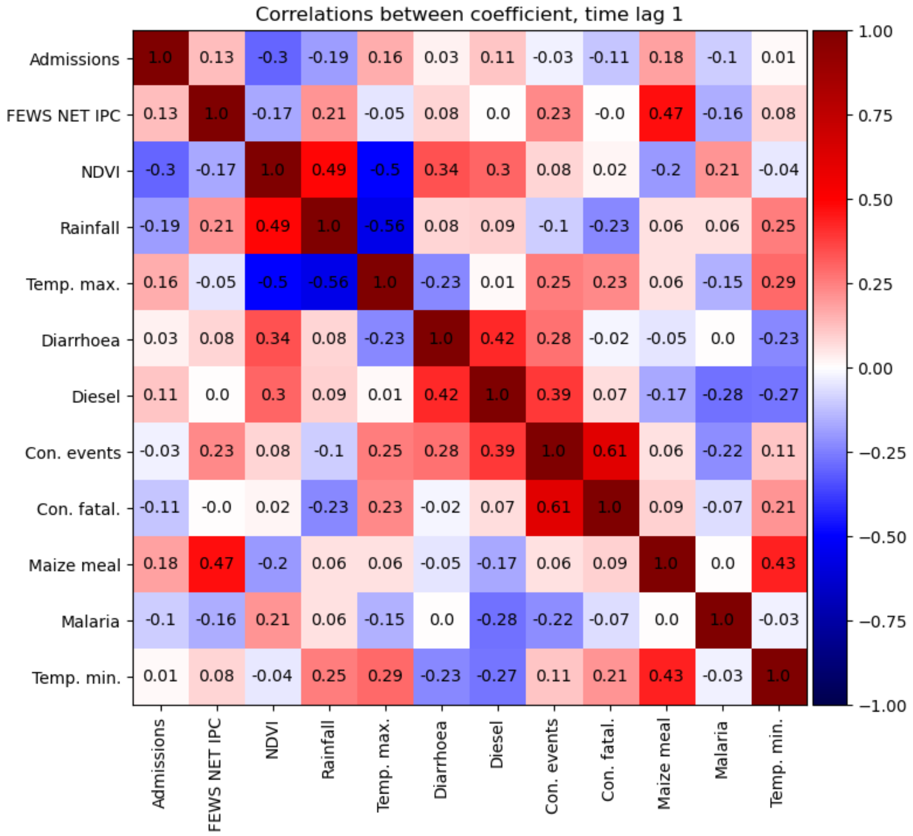


Figure S21: Correlations between coefficients for a forecast horizon of one month.

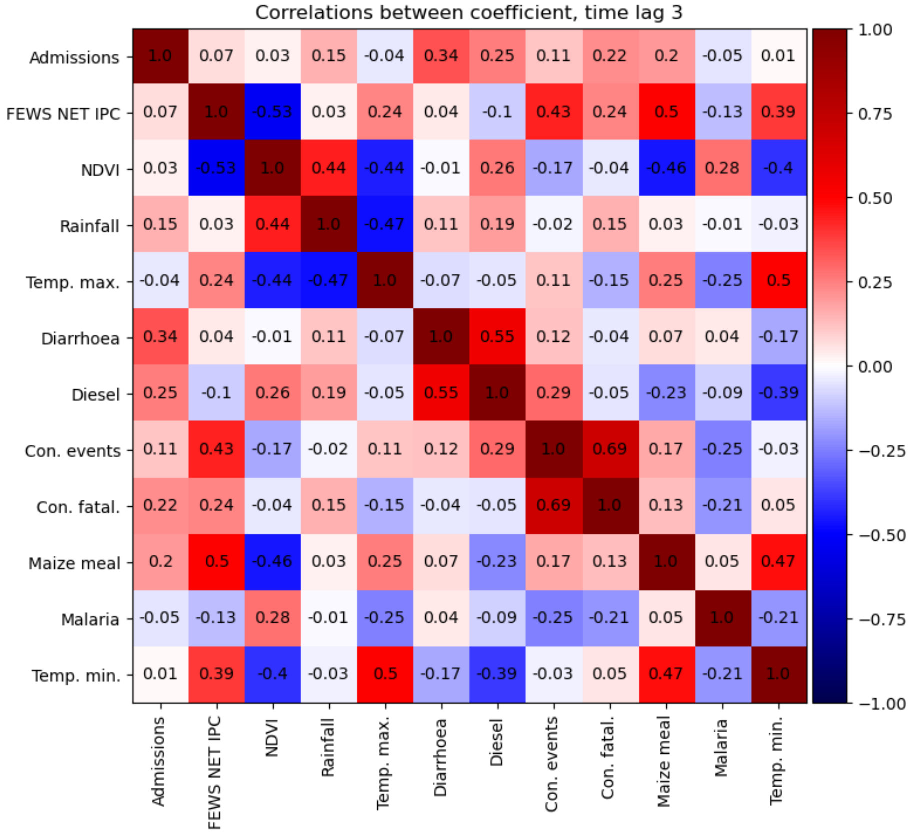


Figure S22: Correlations between coefficients for a forecast horizon of three month.

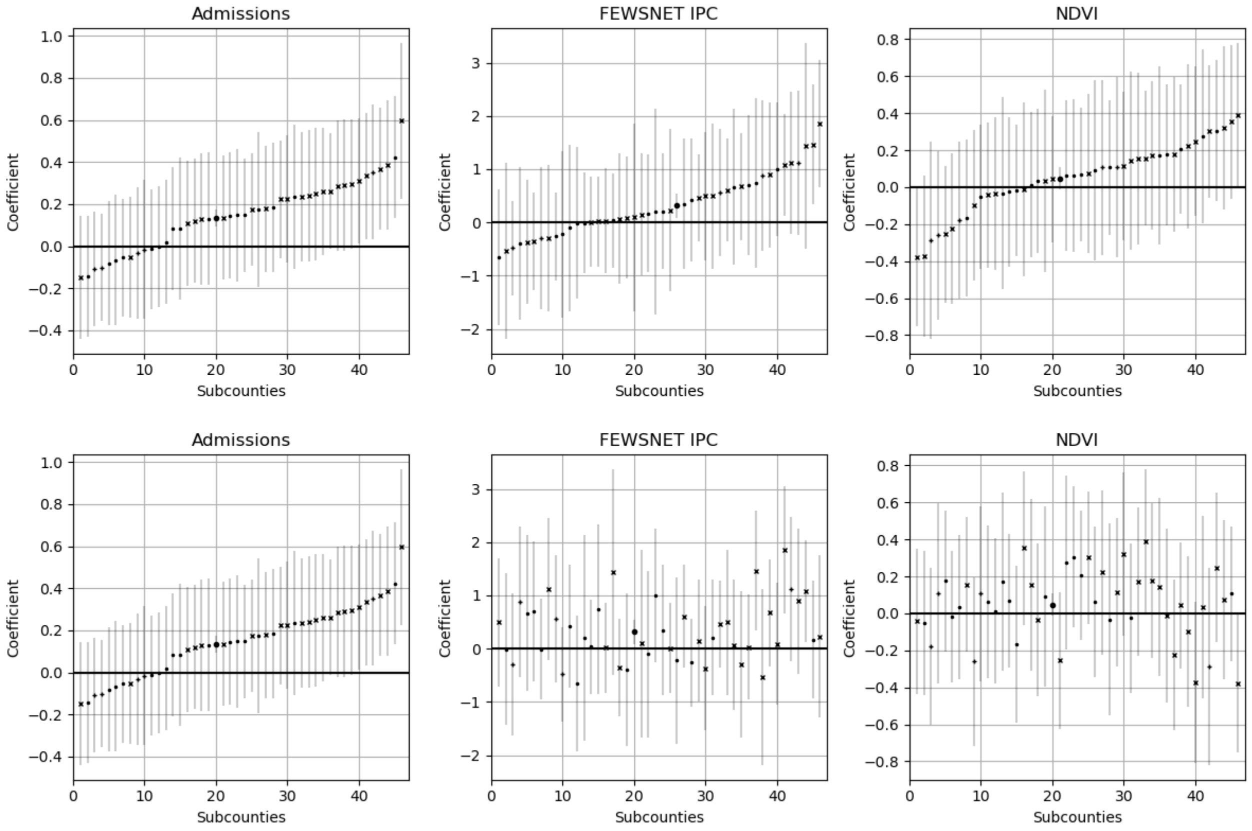


Figure S23: Top) Coefficients by subcounty for multivariable regression with time lags included. The coefficients have been sorted separately for each variable. Bottom) Coefficients by subcounty for univariate regression. The coefficients have been sorted by the same ordering as for the admissions coefficients.

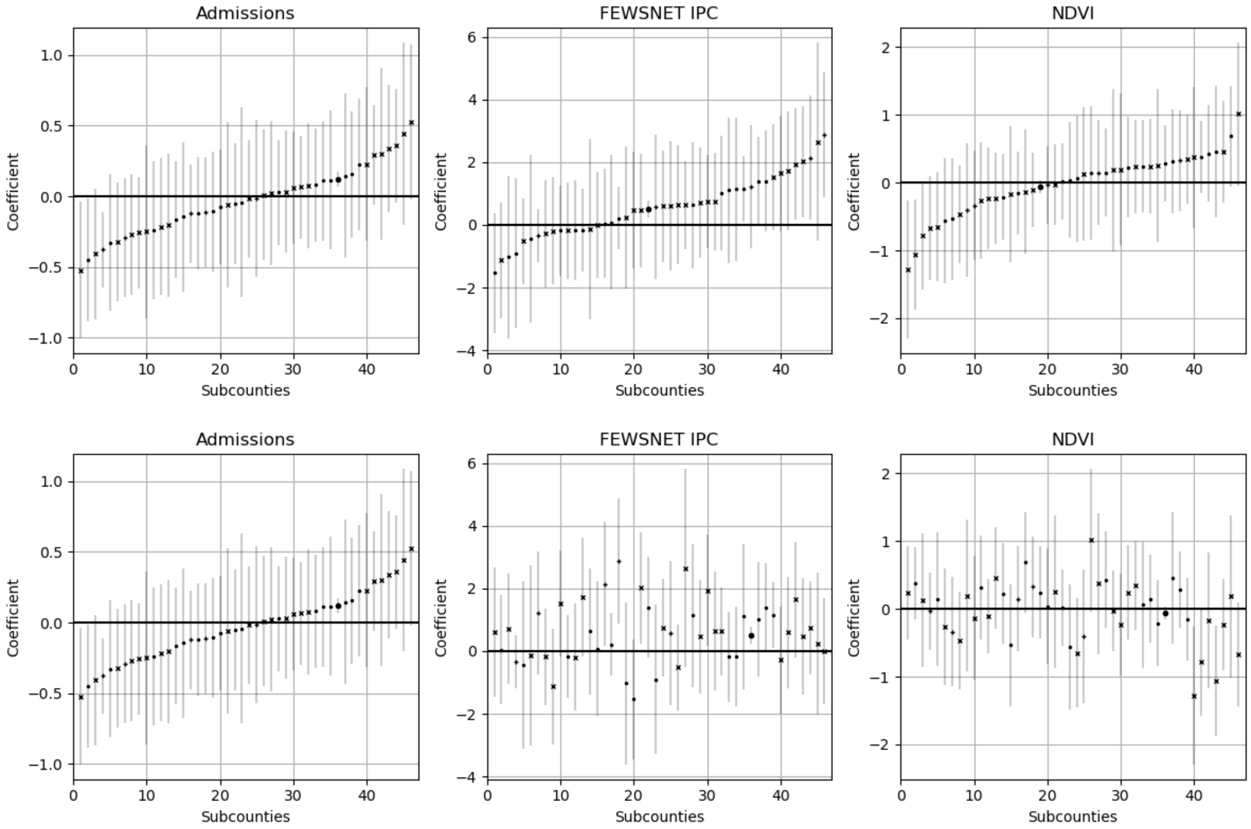


Figure S24: Top) Coefficients by subcounty for multivariable regression with all variables included. The coefficients have been sorted separately for each variable. Bottom) Coefficients by subcounty for univariate regression. The coefficients have been sorted by the same ordering as for the admissions coefficients.

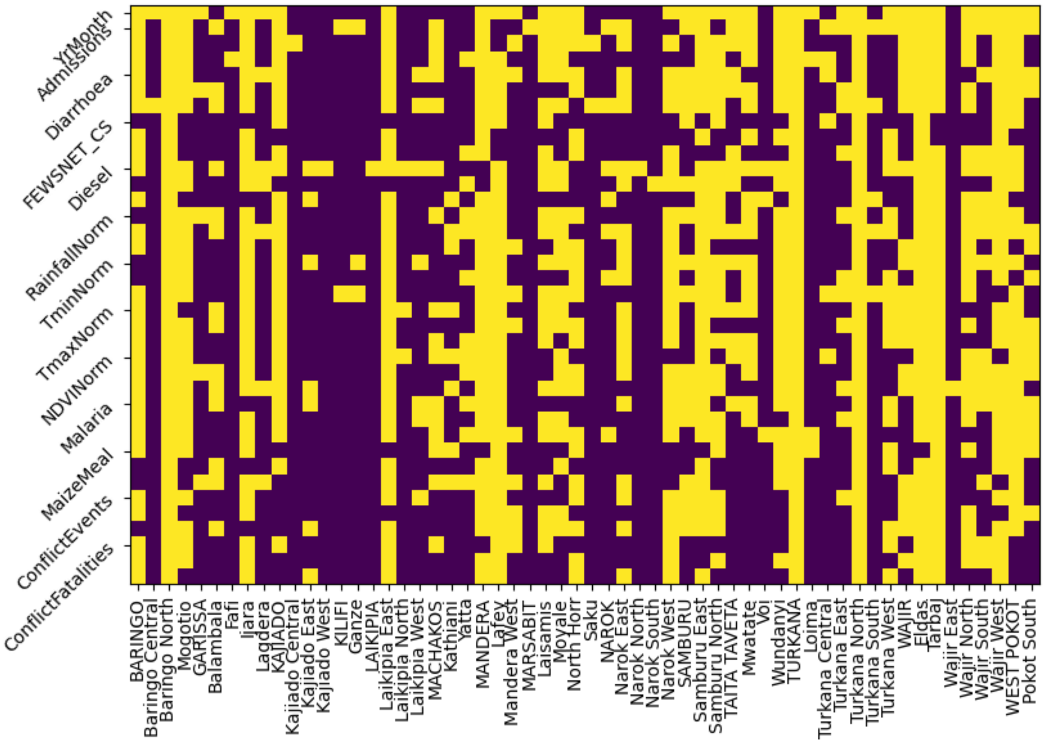


Figure S25: XGBoost feature importance at a forecast horizon of one months demonstrated with retained features (denoted as yellow in the plot) after applying feature selection through cross-validation. There are three squares associated with each variable for the time lags of 0, 1, 2 respectively. On the x-axis capitalised names are counties and those are trained with all the county data. The lower case names following each county name are the subcounties associated with the preceding county. For example BARINGO is a county and Baringo Central, Baringo North and Mogotio are all subcounties within Baringo county.

### 2.3 Temporal concept drift

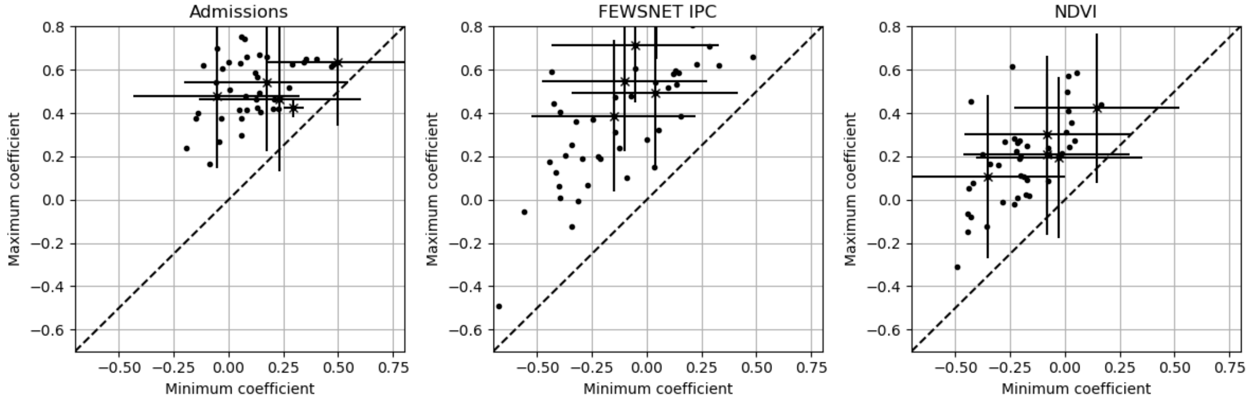


Figure S26: The maximum versus minimum coefficients for three variables (admissions, FEWSNET IPC, NDVI) all at a time-lag of one month when using a 30-month sliding window to train the univariable model.

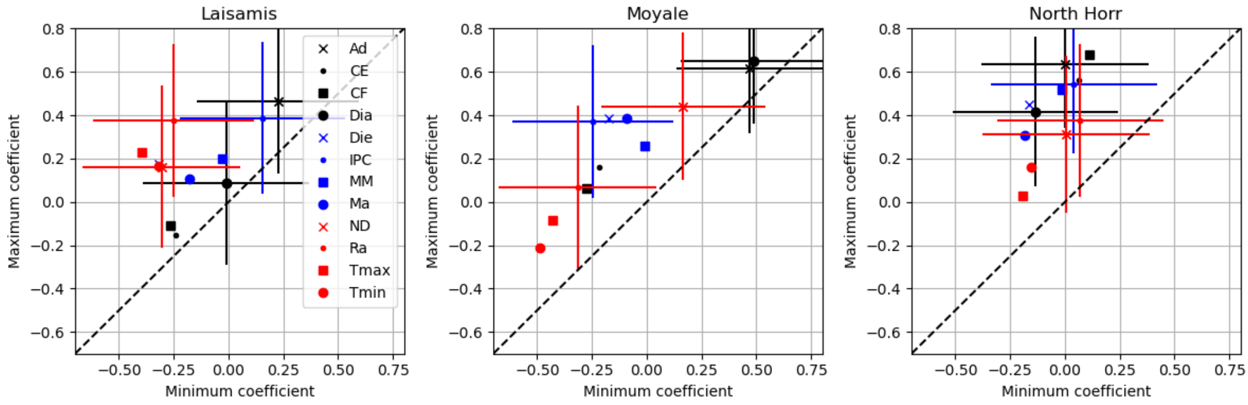


Figure S27: Maximum versus minimum coefficients for all variables at a forecast horizon of one month for an example three subcounties from the county of Marsabit.

### 2.4 Model predictions

Table S2: Table of results for forecast horizon of 1 month. Results for the elastic net (EN) and XGBoost (XGB) are shown as are baselines for average per month (Av. mon.), averaged overall (Av. ov.), the average of the last three months (Av. 3), and the last value (Last). For the models results are shown for different inputs (labelled as In.) with all (A) and autocorrelation alone (AC) shown. Also for the models the training approach using either standard training (St.) or ongoing (On.) is also shown.

| Model | In. | T. | All | Subcounty | County | Livelihood |
|---|---|---|---|---|---|---|
| EN | A | St. | 1.07 (0.96-1.17) | 1.26 (1.12-1.42) | 1.23 (1.08-1.38) | 1.08 (0.97-1.22) |
| EN | A | On. | 1.0 (0.92-1.08) | 1.1 (1.0-1.2) | 1.07 (0.99-1.17) | 1.0 (0.92-1.08) |
| EN | AC | St. | 1.02 (0.93-1.13) | 1.14 (1.0-1.3) | 1.1 (0.96-1.27) | 1.03 (0.93-1.15) |
| EN | AC | On. | 0.96 (0.87-1.05) | 1.04 (0.95-1.15) | 1.0 (0.91-1.09) | 0.96 (0.87-1.06) |
| XGB | A | St. | 1.39 (1.29-1.5) | 1.19 (1.08-1.31) | 1.28 (1.17-1.4) | 1.25 (1.15-1.37) |
| XGB | A | On. | 1.13 (1.05-1.22) | 1.15 (1.06-1.24) | 1.16 (1.08-1.25) | 1.15 (1.06-1.24) |
| XGB | AC | St. | 1.08 (0.96-1.22) | 1.08 (0.94-1.22) | 1.18 (1.06-1.32) | 1.09 (0.96-1.23) |
| XGB | AC | On. | 0.99 (0.9-1.1) | 0.99 (0.89-1.1) | 0.99 (0.89-1.09) | 1.0 (0.9-1.09) |
| Av. mo. | - | - | 1.26 (1.1-1.41) | 1.26 (1.12-1.42) | 1.26 (1.11-1.42) | 1.26 (1.11-1.41) |
| Av. ov. | - | - | 1.23 (1.09-1.39) | 1.23 (1.1-1.38) | 1.23 (1.09-1.39) | 1.23 (1.1-1.38) |
| Av. 3 | - | - | 1.18 (1.05-1.32) | 1.12 (1.01-1.23) | 1.24 (1.12-1.38) | 1.18 (1.07-1.31) |
| Last | - | - | 1.17 (1.04-1.3) | 1.17 (1.06-1.29) | 1.24 (1.12-1.36) | 1.17 (1.04-1.3) |

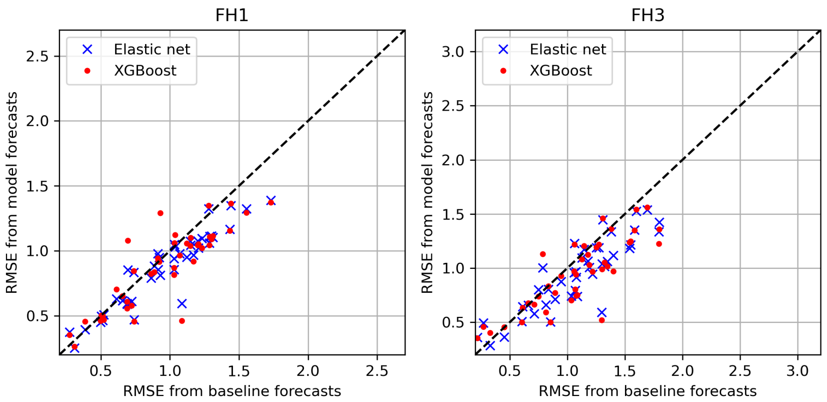


Figure S28: The RMSE per subcounty for model forecasts compared to baseline forecasts for elastic net (blue cross) and XGBoost (red dot). Left) Forecast horizon of 1 month (FH1). Right) Forecast horizon of 3 months (FH3).

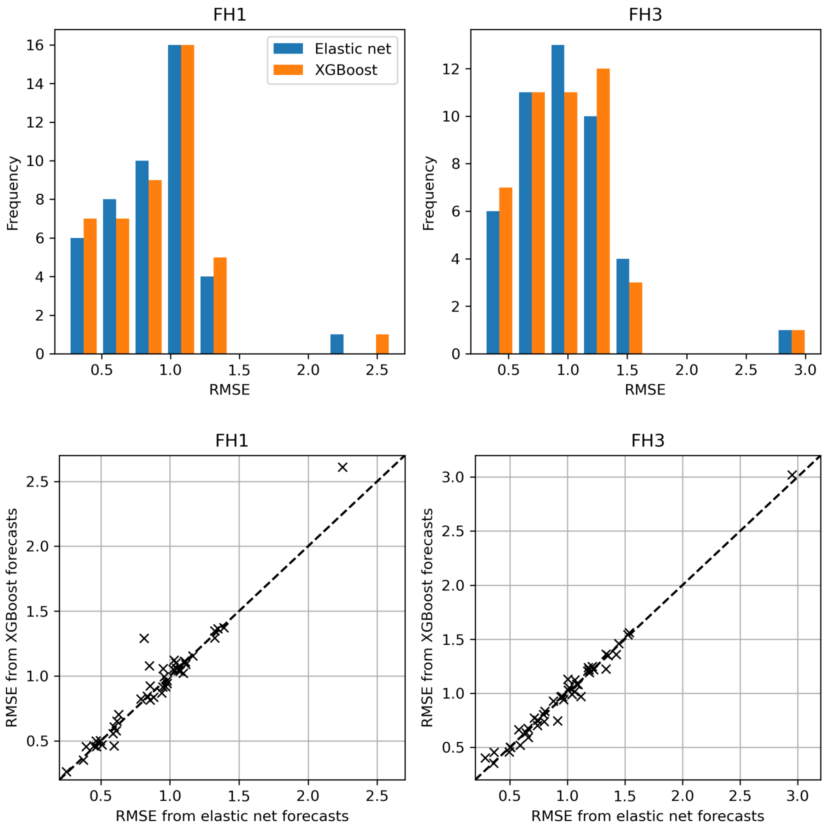


Figure S29: Comparisons of elastic net and XGBoost RMSE metrics per subcounty, both for just the autocorrelation. Top) Histograms of the RMSEs by subcounty for the models. Bottom) Direct comparisons of the RMSEs by subcounty for the two models. Left) Forecast horizon of 1 month (FH1). Right) Forecast horizon of 3 months (FH3).

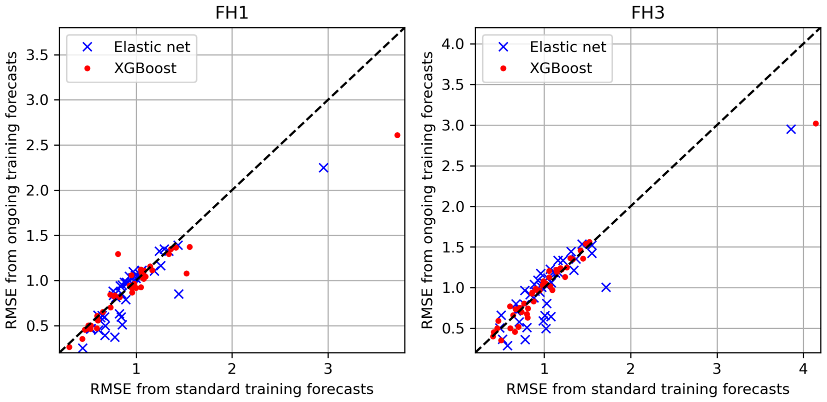


Figure S30: Comparison of the RMSE for forecasts when the models are trained in an ongoing (y-axis) or a standard (x-axis) approach, both for just the autocorrelation. Left) Forecast horizon of 1 month (FH1). Right) Forecast horizon of 3 months (FH3).

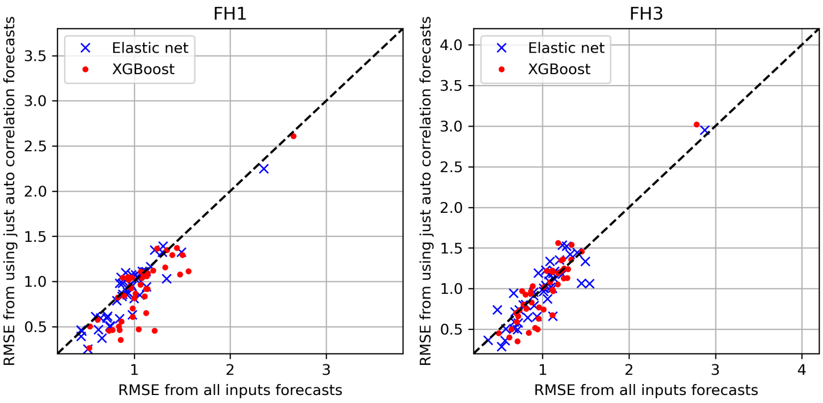


Figure S31: Comparison of the RMSE for forecasts when the models are trained using all the inputs (x-axis) or using just autocorrelation (y-axis), both trained in an ongoing manner. Left) Forecast horizon of 1 month (FH1). Right) Forecast horizon of 3 months (FH3).
